# Combined Metabolic Activators Reduces Liver Fat in Nonalcoholic Fatty Liver Disease Patients

**DOI:** 10.1101/2021.05.20.21257480

**Authors:** Mujdat Zeybel, Ozlem Altay, Muhammad Arif, Xiangyu Li, Hong Yang, Claudia Fredolini, Murat Akyildiz, Burcin Saglam, Mehmet Gokhan Gonenli, Dilek Ural, Woonghee Kim, Jochen M. Schwenk, Cheng Zhang, Saeed Shoaie, Jens Nielsen, Mathias Uhlén, Jan Borén, Adil Mardinoglu

**Author notes:** **Clinical trial registration number:** NCT04330326 (https://clinicaltrials.gov/).

## Abstract

Nonalcoholic fatty liver disease (NAFLD) refers to excess fat accumulation in the liver. In animal experiments and human kinetic study, we found that administration of combined metabolic activators (CMA) promotes the oxidation of fat, attenuates the resulting oxidative stress, activates mitochondria and eventually removes excess fat from the liver. Here, we tested the safety and efficacy of CMA in NAFLD patients in a placebo-controlled 10-week study. We found that CMA significantly decreased hepatic steatosis and levels of aspartate aminotransferase, alanine aminotransferase, uric acid, and creatinine, whereas found no differences on these variables in the placebo group after adjustment for weight loss. By integrating clinical data with plasma metabolomics and inflammatory proteomics as well as oral and gut metagenomics data, we revealed the underlying molecular mechanisms associated with the reduced hepatic fat and inflammation in NAFLD patients and identified the key players involved in the host-microbiome interactions. In conclusion, we observed that CMA can be used develop a pharmacological treatment strategy in NAFLD patients.

## INTRODUCTION

Nonalcoholic fatty liver disease (NAFLD), defined as the hepatic fat accumulation of ≥5% unrelated to alcohol consumption and other liver diseases, comprises pathologies that include hepatic steatosis, steatohepatitis, and hepatic fibrosis and cirrhosis (Anstee *et al*, 2019). NAFLD is closely associated with insulin resistance and metabolic syndrome. Due to the rapid rise in the prevalence of obesity and diabetes; it is the leading cause of chronic liver disease (Estes *et al*, 2018; Younossi *et al*, 2018). Globally, at least one in four people have hepatic steatosis (Abeysekera *et al*, 2020; Bugianesi, 2020).

Current management strategies, including lifestyle modification, increased physical activity, and dietary intervention, have limited adherence and minimal prolonged success (Alferink *et al*, 2019; Romero-Gómez *et al*, 2017; Sanyal, 2019). No drugs have been approved to treat NAFLD, and effective treatment options with long-term safety are urgently required. Although research is paving the way for the development of therapeutics, the results of early clinical trials of drugs targeting single pathways have been mostly unsuccessful. Combining compounds that reduce lipid accumulation and hepatocellular injury has been proposed as a more suitable therapeutic strategy for this complex disease (Barbier-Torres *et al*, 2020; Barbier-Torres *et al*, 2017; Ertunc & Hotamisligil, 2016). Targeting multiple pathways is more likely to translate into successful outcomes (Friedman *et al*, 2018; Mardinoglu *et al*, 2018a; Mardinoglu *et al*, 2019).

A key mechanism in the pathogenesis of NAFLD is inadequate removal of hepatic fat by fatty acid oxidation. Our previous work combining in-depth multi-omics profiling and hepatocyte-specific integrated networks identified three landmark metabolic features of hepatic steatosis: limited serine availability, reduced *de novo* glutathione (GSH) synthesis, and altered nicotinamide adenine dinucleotide (NAD+) metabolism (Lee *et al*, 2016; Mardinoglu *et al*, 2014; Mardinoglu *et al*, 2017).

We hypothesized that NAFLD could be treated with combined metabolic activators (CMA), including L-carnitine tartrate to facilitate mitochondrial fatty acid uptake from cytosol; the NAD^+^ precursor nicotinamide riboside to induce hepatic mitochondrial β-oxidation and facilitate fatty acid transfer through the mitochondrial membrane, and the potent glutathione precursors L-serine and N-acetyl-l-cysteine to reduce oxidative stress (Mardinoglu *et al*, 2018b). We further hypothesized that administration of these metabolic activators would promote mitochondrial fatty acid uptake and oxidation and reduce hepatic fat and inflammation. In animal toxicology studies and a human calibration study for CMA, we found that the metabolic activators were well tolerated and increased the activators’ plasma levels and their associated metabolites. Additionally, administration of CMA effectively increased fatty acid oxidation and *de novo* glutathione generation, as judged by metabolomic and proteomic profiling (Zhang *et al*, 2020). In this placebo-controlled phase 2 study, we tested our hypotheses and the efficacy and safety of CMA in NAFLD patients by integrating clinical data with plasma metabolomics and inflammatory proteomics as well as oral and gut metagenomics.

## RESULTS

### Patient Characteristics

Of 56 patients screened for the trial, 32 met the eligibility criteria (see Methods and Supplementary Appendix) and were randomly assigned to receive treatment or placebo. Twenty-four patients were excluded according to the study protocol (Datasets S1, S2 and S3), and one eligible patient moved to another city before the randomization. Of the remaining 31 eligible patients, 20 were randomly assigned to the CMA group and 11 to the placebo group (Figure 1A). One patient was excluded from analysis as a result of the COVID-19 lockdown. The 30 remaining patients completed the study (Figure 1A). However, because of the COVID-19 lockdown, eight were unable to visit the Day 14 visit’s trial site but completed the final visit on Day 70. The patients’ mean age was 40.4 years (25–63 years), and 77.4% were men (Table 1). Baseline demographic and clinical characteristics did not differ between groups (Table 1, Dataset S4).

**TABLE 1.** Baseline demographics of the study population and summary of significantly different clinical parameters in the CMA and placebo groups after weight loss adjustment.

|  | **CMA group (n = 20)** |  |  |  |  | **Placebo group (n = 11)** |  |  |  |
| --- | --- | --- | --- | --- | --- | --- | --- | --- | --- |
| Age, yr (range) | 40.9 (25–62) |  |  |  |  | 39.5 (27–63) |  |  |  |
| Male, n | 16 (80.0%) |  |  |  |  | 8 (72.7%) |  |  |  |
| Female, n | 4 (20.0%) |  |  |  |  | 3 (27.3%) |  |  |  |
|  | **Day 14 vs Day 0** |  | **Day 70 vs Day 0** |  |  | **Day 14 vs Day 0** |  | **Day 70 vs Day 0** |  |
|  | **FC** | **p.adj value\*** | **FC** | **p.adj value\*** |  | **FC** | **p.adj value\*** | **FC** | **p.adj value\*** |
| Hepatic fat (%) | 0.895 | 0.096 | 0.887 | **0.033** |  | 0.963 | 0.968 | 0.969 | 0.825 |
| ALT (IU/L) | 0.723 | **0.0015** | 0.630 | **5.75e-07** |  | 0.827 | 0.458 | 0.767 | 0.062 |
| AST (IU/L) | 0.848 | 0.061 | 0.735 | **1.58e-05** |  | 0.915 | 0.796 | 0.822 | 0.059 |
| Uric acid (mg/dL) | 0.886 | **0.003** | 0.853 | **1.12e-05** |  | 0.906 | 0.556 | 0.912 | 0.531 |
| Creatinine (mg/dL) | 0.965 | 0.209 | 0.925 | **0.0007** |  | 1.009 | 0.968 | 0.996 | 0.961 |

FC, fold change.
\*Adjusted for weight loss.

**Figure 1.**
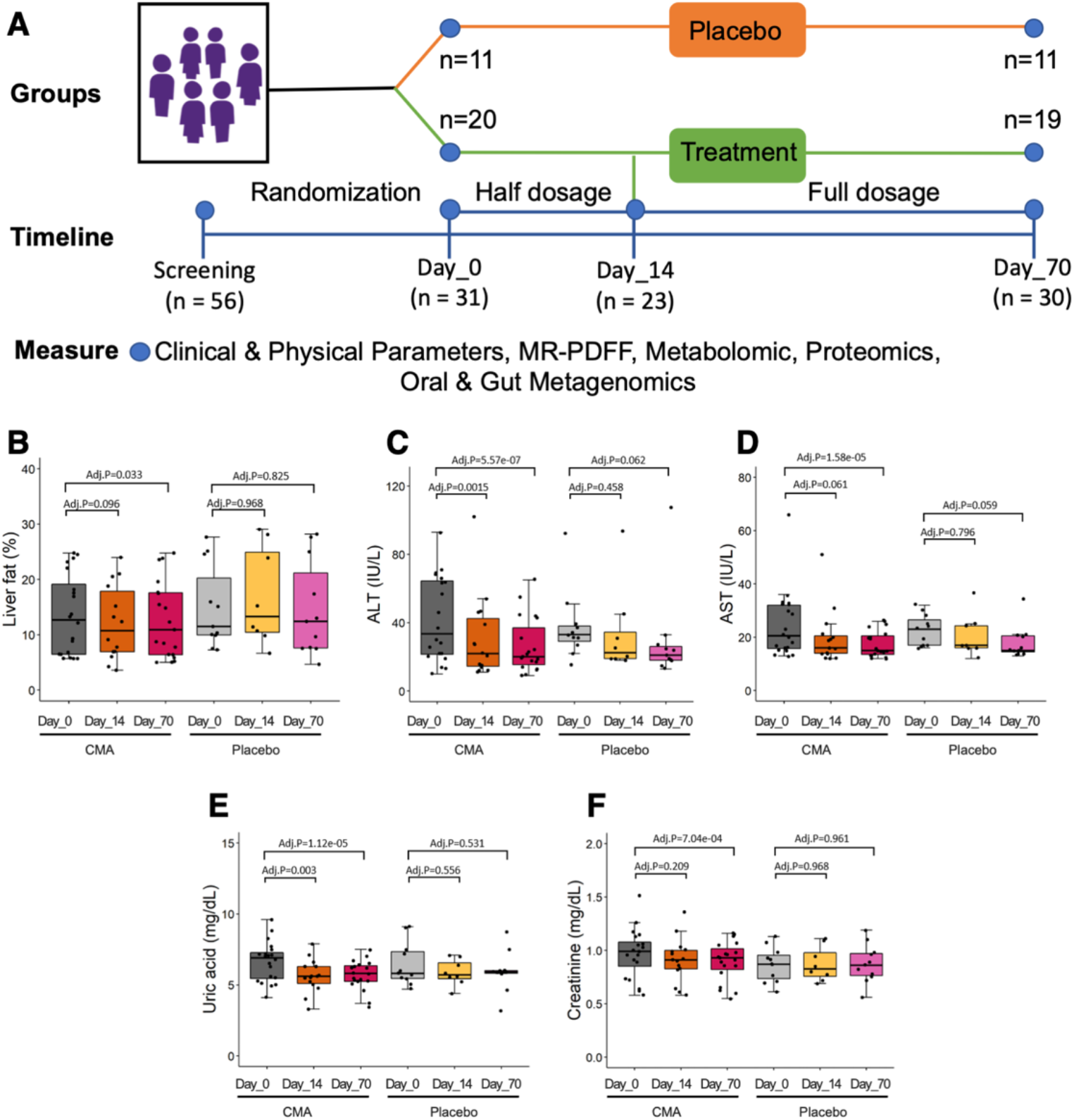
CMA Improves Liver Fat and Clinical Parameters. (A) Study design for testing the effects of CMA in NAFLD patients. Differences in clinical variables including (B) liver fat, plasma levels of (C) ALT, (D) AST, (E) uric acid and (F) creatinine are presented in the CMA and placebo groups on Days 0, 14 and 70 after weight loss adjustment. Adj.P indicates p value after weight loss adjustment. Statistical significance is defined based on paired Student’s t test. p< 0.05.

### CMA Decreases Hepatic Steatosis and Improves Clinical Parameters

Adherence to the treatment regimen was more than 95% and did not differ between groups. The primary outcome variable, hepatic fat content as judged from the proton density fat fraction estimated by magnetic resonance imaging (MRI-PDFF), was lower on Day 70 than on Day 0 in the CMA group (10%, p = 0.028) but not in the placebo group (Figure 1B, Dataset S3 & S4). After adjustment for weight loss, this difference remained significant in the CMA group (p.adj = 0.033) (Figure 1, Table 1).

The serum level of alanine aminotransferase (ALT) was lower on Day 70 in both in the CMA group (39%, p = 0.0003 vs Day 0) and placebo group (22%, p = 0.025); however, after weight loss adjustment, the difference remained significant only in the CMA group (p.adj = 5.75e-07, Table 1). On Day 14, the ALT level after weight loss adjustment was significantly lower only in the CMA group (24%, p.adj = 0.0015) (Figure 1C, Table 1, Dataset S4). Similarly, the serum level of aspartate aminotransferase (AST) on Day 70 vs Day 0 was lower in both the CMA group (30%, p = 0.004) and the placebo group (20%, p = 0.009); however, after weight loss adjustment, the AST level was significantly lower only in the CMA group (p.adj = 1.58e-05) (Figure 1D, Table 1, Dataset S4).

The serum uric acid level was lower in the CMA group on both Day 14 (12%, p.adj = 0.003) and Day 70 (15%, p.adj = 1.12e-05), both before and after weight loss adjustment (Figure 1E, Table 1, Dataset S4). However, the creatinine level on Day 70 was significantly decreased only in the CMA group (8%, p.adj=.0007), both before and after weight loss adjustment (Figure 1F, Table 1, Dataset S4). The serum levels of uric acid and creatinine did not change in the placebo group at either time point (Table 1, Dataset S4). Systolic blood pressure on Day 70 was 5% lower in the CMA group (p = 0.023), whereas in the placebo group, it was unchanged and diastolic blood pressure had increased (p = 0.038) (Dataset S4).

### CMA Alters Plasma Metabolites

To characterize the patients and reveal the underlying molecular mechanisms associated with the decrease in liver fat and improvement in clinical parameters in the CMA group, we generated untargeted metabolomics data from plasma samples and measured the levels of 1032 metabolites (Dataset S5). After excluding metabolites with missing values in >50% of samples, 929 metabolites were analyzed (Dataset S5). Metabolites, whose levels on Day 70 and Day 14 vs Day 0 differed significantly both before and after weight loss adjustment, are listed in Dataset S6; those that differed between groups are listed in Dataset S7.

Plasma levels of serine, nicotinamide, and carnitine, which are directly associated with CMA, and of metabolites indirectly associated with CMA were significantly higher on Day 70 than on Day 0 in the CMA group (Figure 2A, Dataset S6), and those levels were significantly higher than in the placebo group (Dataset S7). Specifically, the plasma levels of N1-methyl-4-pyridone-3-carboxamide, N1-methyl-2-pyridone-5-carboxamide, and 1-methylnicotinamide (associated with NR and NAD^+^ metabolism); of N-acetylglycine, N-palmitoylserine, N-oleoylserine, and N-stearoylserine (associated with serine and glycine metabolism); and of deoxycarnitine, acetylcarnitine, and butyrylcarnitine (associated with carnitine metabolism) were significantly higher on Day 70 than on Day 0 in the CMA group (Dataset S6). On the other hand, we found that the plasma level of cysteine associated with NAC showed tendency (p=.052) to be down-regulated on Day 70 vs Day 0 in the CMA group (Figure 2A, Dataset S6). Hence, we observed that NAC has different plasma kinetics compared to the other three metabolic activators. We studied the association of the plasma level of individual metabolic activators with the plasma level of the metabolites (Figure 2B, Dataset S8). We provided a mechanistic explanation for the alterations in the plasma level metabolites other than administered metabolic activators.

**Figure 2.**
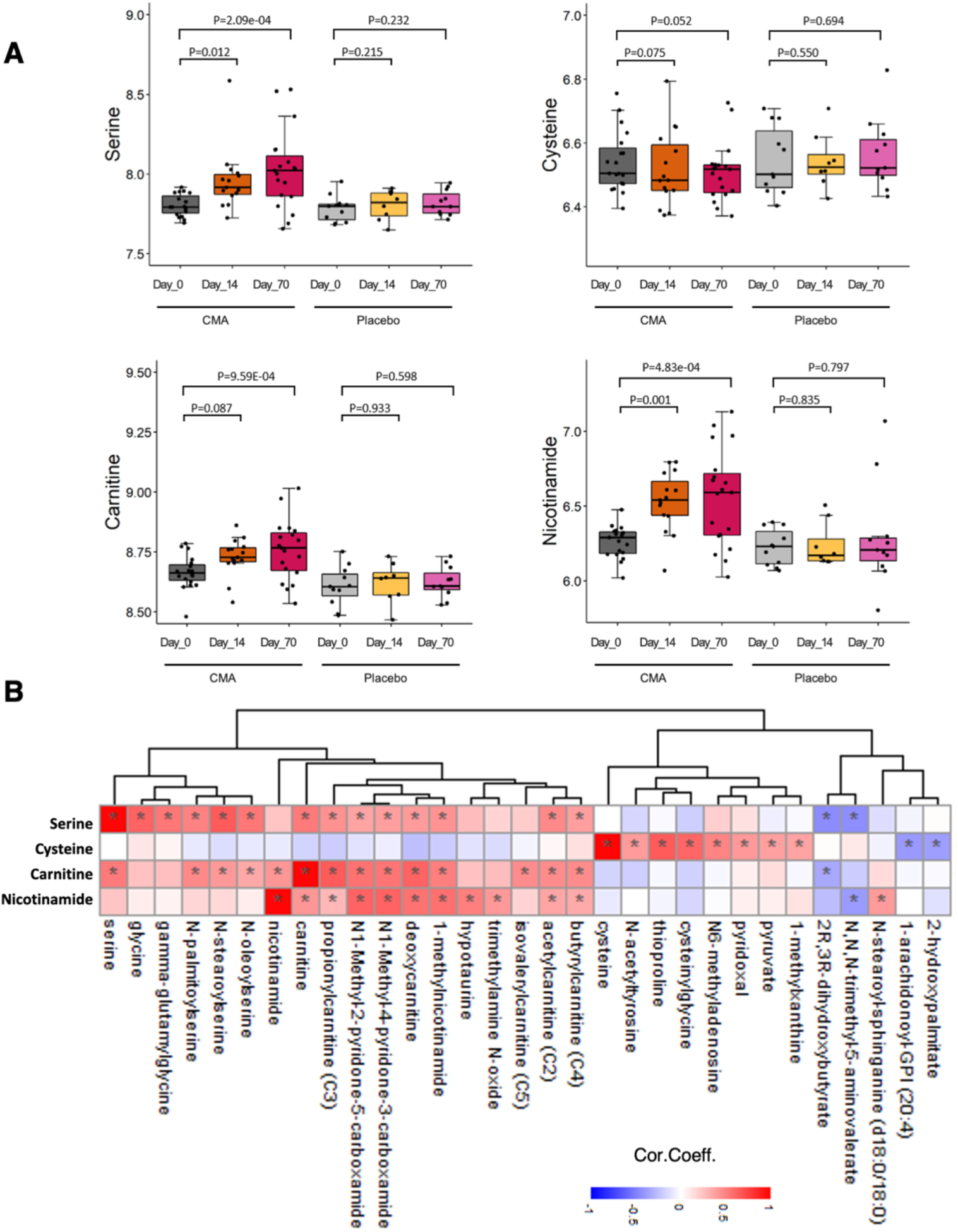
Supplementation Improves the Plasma Level of Metabolic Activators. (A) Differences in the plasma levels of individual CMA including serine, carnitine, cysteine and nicotinamide are shown in the CMA and placebo groups on Days 0, 14 and 70. The y axis is log10 transformation. B) Associations between the plasma level of individual CMA and the 10 most significantly correlated plasma metabolites are presented. Asterisks indicate statistical significance (FDR < 0.05) based on Spearman correlation analysis. Only metabolites detected in more than 50% of samples are included in the analysis. Cor.Coeff: Correlation coefficient

We found that 110 metabolites differed significantly on Day 70 vs Day 0 (p ≤ 0.05) in the CMA group; 44 metabolites were involved in lipid metabolism (Figure S1), and 66 metabolites were involved in amino acids and other parts of the metabolism (Figure 3A, Dataset S6). Of the 66 metabolites that were significantly different on Day 70 vs Day 0 in the CMA group, 4 of them were significantly different in both groups (Figure 3B), 62 of them were significantly different in only CMA group (Figure 3C), and 27 of them were significantly different only in the placebo group (Figure 3D). Previously, we found that the increased plasma levels of kynurenine and kynurenate were associated with high hepatic fat (Mardinoglu *et al*., 2018b). In a one-day clinical study of the acute effect of CMA on plasma metabolite levels, we found that kynurenine and kynurenate levels were significantly decreased (Zhang *et al*., 2020). In the current study, plasma levels of kynurenate (Figure 3B) and kynurenine (Figure 3C) was significantly lower on Day 70 vs Day 0 in the CMA group both before and after weight loss adjustment (Dataset S6). On the other hand, the kynurenate plasma level was significantly increased on Day 70 vs Day 0 in the placebo group (Figure 3B). Moreover, we found that the plasma level of 3-amino-2-piperidone associated with the urea cycle; arginine and proline metabolism were significantly decreased on Day 70 vs Day 0 in the CMA group, whereas its plasma level was significantly increased in the placebo group (Figure 3B).

**Figure 3.**
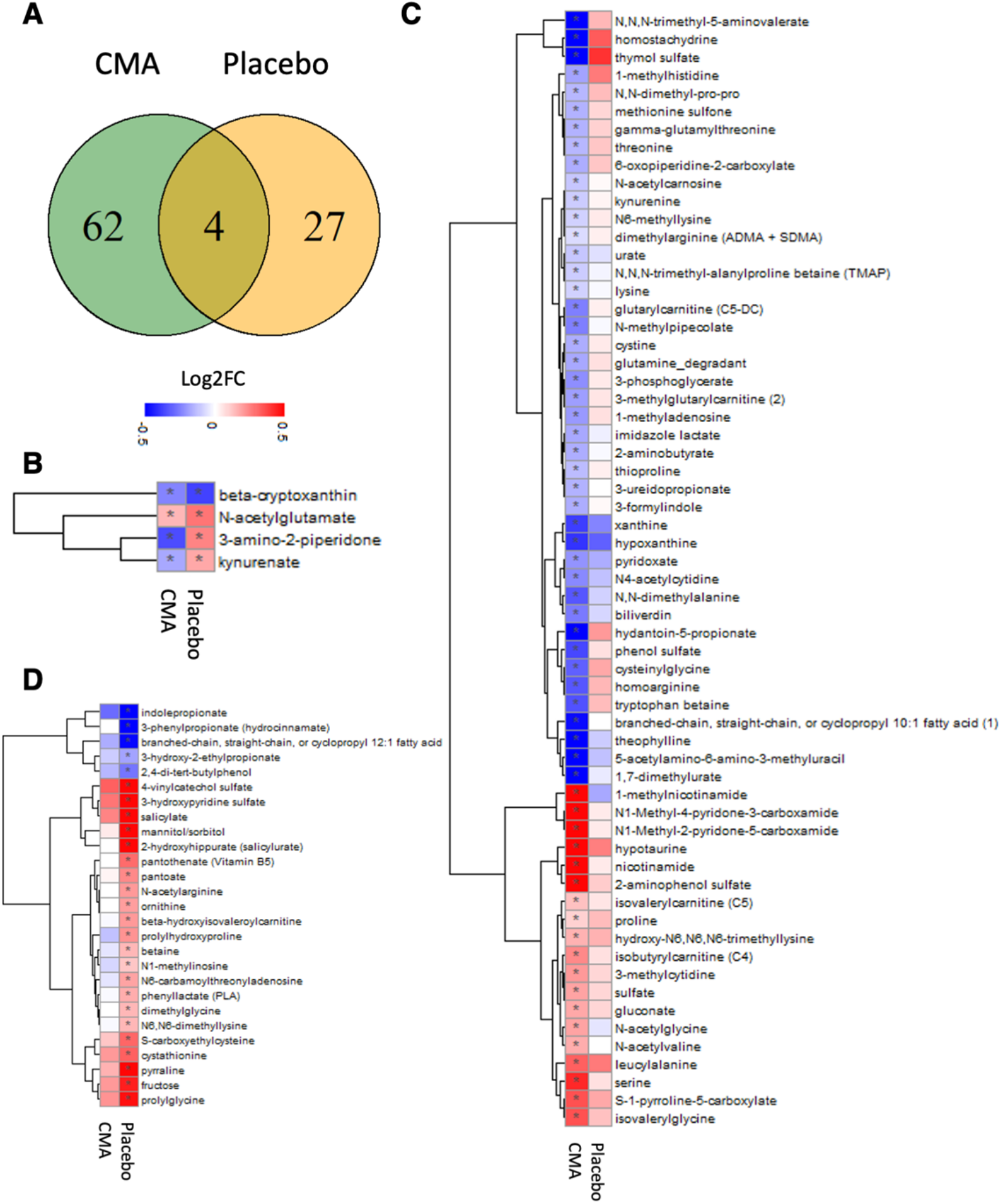
CMA Alters Plasma Metabolite Levels. Plasma level of metabolites (other than lipids) that are significantly different between Day 70 vs Day 0 in the CMA and placebo groups after weight loss adjustment are presented. A) Venn-diagram represents the number of significantly different metabolites (other than lipids) on Day 70 vs Day 0 in the CMA and placebo groups. Statistical significance is defined based on paired Student’s t test. p< 0.05. Association between the plasma level of significantly different metabolites on Day 70 vs Day 0 B) in both (n=4); C) only in CMA (n=62) and D) only in placebo (n=27) groups are shown. Heatmap shows log2FC values of metabolites between Day 70 vs Day 0. Asterisks indicate statistical significance based on paired Student’s t test. p< 0.05. Log2FC: log2(fold change).

Our analysis also revealed decreased metabolism of purine and xanthine in the CMA group on Day 70. Plasma levels of urate, xanthine, hypoxanthine 5-acetylamino-6-amino-3-methyluracil, 1,7-dimethylurate, and theophylline were significantly reduced in the CMA group on Day 70 (Figure 3C, Dataset S6). Consistent with these findings, plasma uric acid levels were significant decreased in this group. N-trimethyl-5-aminovalerate (TMAVA) was the most significantly reduced metabolite in the CMA group on Day 70, both before and after weight loss adjustment (Figure 3C, Dataset S6) and was significantly lower than in the placebo group (Dataset S7). TMAVA is linked to intestinal microbes and associated with lysine metabolism and is one of the best predictors of microalbuminuria (Haukka *et al*, 2018). In a previous study, the plasma level of TMAVA was significantly increased in NAFLD patients (Zhao *et al*, 2020). *Enterococcus faecalis* and *Pseudomonas aeruginosa* are responsible for metabolizing trimethyllysine to TMAVA, which was further modulated by antibiotic treatment in a mouse model (Zhao *et al*., 2020). We also found that the plasma level of N,N,N-trimethyl-alanylproline betaine (TMAP) was significantly downregulated in the CMA group on Day 70, even after weight loss adjustment (Figure 3C, Datasets S6), as were creatinine levels (Figure 1F, Table 1, Dataset S4). TMAP has been described as a novel potential biomarker of the dialytic clearance that can accurately define kidney function (Velenosi *et al*, 2019).

CMA treatment correlated significantly with the plasma levels of serine, glycine, gamma-glutamylglycine, carnitine, TMAVA, 1-methylnicotinamide, N1-methyl-4-pyridone-3-carboxamide, and N1-methyl-2-pyridone-5-carboxamide (Figure 2B, Dataset S8). But none of these metabolites correlated significantly with the plasma cysteine level, indicating that cysteine utilization differs from that of other metabolites in the CMA.

### CMA Reduces Inflammation

Plasma levels of 96 inflammatory protein markers were measured with the plasma proteome profiling platform Proximity Extension Assay (PEA) using an inflammation panel quantifying the plasma level of target proteins. After quality control and exclusion of proteins with missing values in more than 50% of samples, 72 proteins were analyzed (Dataset S9). Proteins whose levels differed significantly between the visits in the CMA and placebo groups are listed in Dataset S10. The plasma levels of CD8A, CSF-1, CCL23, FGF-21, and oncostatin-M (OSM) were significantly decreased only in the CMA group (Figure 4A, Dataset S10); however, no significant changes of plasma levels of inflammation-related proteins were found in the placebo group (Dataset S10).

**Figure 4.**
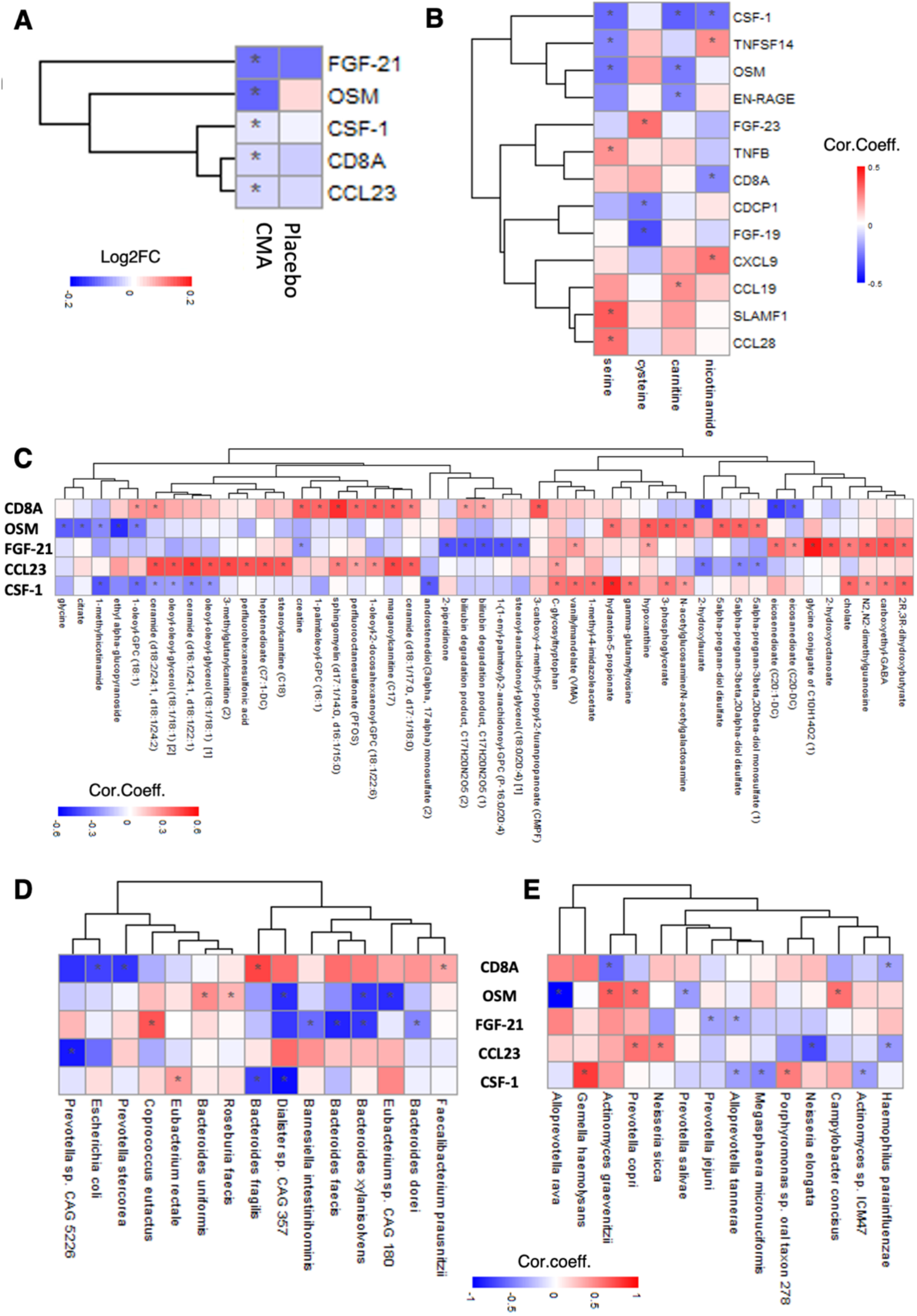
CMA Decreases Plasma Levels of Inflammatory Proteins. **A)** Heatmap shows log2FC based alterations between the significantly different inflammation related proteins on Day 70 vs Day 0 in the CMA and placebo groups. Asterisks indicate statistical significance based on paired Student’s t test. p < 0.05; **B)** Heatmap shows the correlation between the plasma levels of all inflammation related proteins and plasma levels of the individual metabolic activators including serine, cysteine, carnitine and nicotinamide. Asterisks indicate statistical significance based on Spearman correlation analysis. p < 0.05; Heatmap shows the associations between the significantly different inflammation related proteins (CD8A, CSF-1, CCL23, FGF-21 and OSM) **C)** with the 10 most significantly correlated plasma metabolites, **D)** with the abundance of the species in the gut microbiome and **E**) with the abundance of the species in the oral microbiome. Asterisks indicate statistical significance based on Spearman correlation analysis. P < 0.05; Cor.Coeff: Correlation coefficient; Log2FC: log2(fold change).

The plasma level of FGF-21 is increased in NAFLD patients and is a potential diagnostic marker of NAFLD (Rusli *et al*, 2016). The plasma level of FGF-21 correlates positively with high hepatic fat levels in both mice and humans (Dushay *et al*, 2010). The plasma levels of CCL23 and CD8A have also been associated with hepatic steatosis (Marra & Lotersztajn, 2013). We recently found that the plasma levels of CSF-1, OSM, and FGF-21 are significantly associated with hepatic steatosis (Lovric *et al*, 2018). The findings in this study are in agreement with our previous studies. The plasma levels of FGF21 and CCL23 rapidly decreased in our 1-day CMA study after eliminating the effect of the fasting (Zhang *et al*., 2020) and the plasma level of FGF21, CCL23 and CSF-1 had significantly reduced in one-week carbohydrate-restricted diet study (Mardinoglu *et al*., 2018b).

We assessed the associations between the plasma level of the significant proteins and the individual metabolic activators and found that CSF-1 and OSM levels are negatively correlated with carnitine and serine levels (Figure 4B, Dataset S11). Additionally, nicotinamide is also negatively correlated with CSF-1 levels (Figure 4B, Dataset S11).

We finally identified links between proteins whose plasma levels changed significantly and plasma metabolites (Figure 4C, Dataset S12). FGF-21 correlated with the glycine conjugate of C10H14O2, 2-hydroxyoctanoate, and N2,N2-dimethylguanosine and correlated negatively with bilirubin degradation products, 2-piperidinone, and carboxyethyl-GABA.

### CMA alters gut and oral microbiome

The alterations in the gut microbiome have been associated with the NAFLD(Aron-Wisnewsky *et al*, 2020). In our study, we collected feces and saliva samples to study the effect of the CMA on the gut and oral microbiome and revealed the interactions between the host and microbe interactions during the CMA treatment.

We first compared the differences in the species’ abundances between Day 70 vs Day 0 in the CMA and placebo groups in the gut microbiome. We found that abundances of Proteobacteria (*Neisseria flavescens*), Actinobacteria (*Rothia mucilaginosa, Adlercreutzia equolifaciens, Asaccharobacter celatus, Collinsella aerofaciens, Bifidobacterium adolescentis and Actinomyces* sp ICM47) and Firmicutes (*Streptococcus mitis, Streptococcus sanguinis, Streptococcus parasanguinis, Roseburia faecis, Roseburia hominis and Eubacterium hallii*) were significantly decreased. However, in the placebo group species belong to Firmicutes (*Firmicutes bacterium* CAG 83 and *Oscillibacter* sp 57 20), and *Bacteroides vulgatus* were significantly reduced (Figure 5A, Dataset S16 & S17). Numerous studies indicated that Actinobacteria and Proteobacteria abundance was increased in NAFLD compared to healthy controls, at the level of phylum(Guohong *et al*, 2019). Furthermore, increased Firmicutes-to-Bacteroidetes ratio has been stated as a feature of obesity-related NAFLD, but subsequent studies have shown inconsistent results, which still needs further research(Porras *et al*, 2018).

**5.**
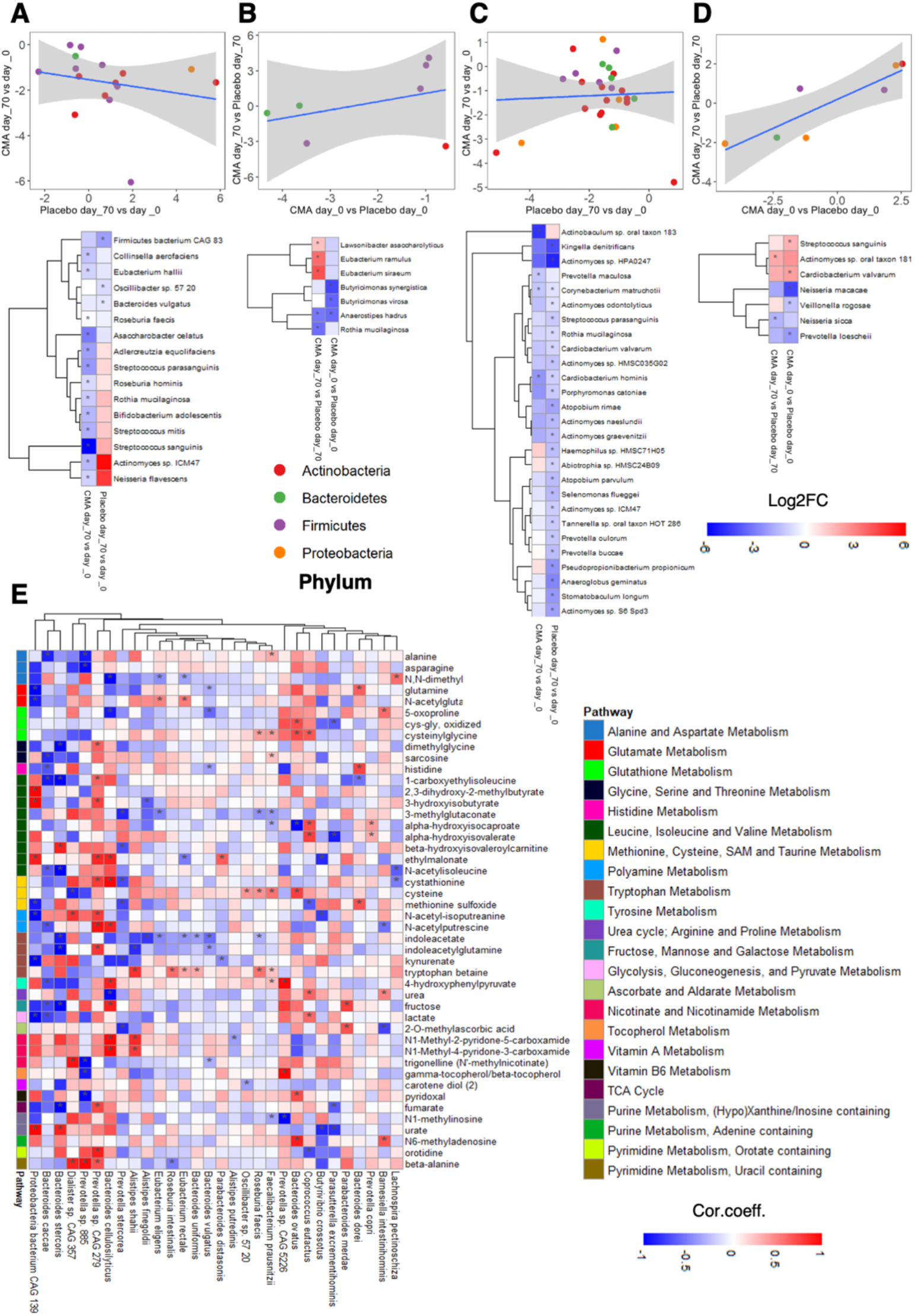
CMA alters gut and oral microbiome. Scatter plot with linear regression line and heatmap show log2FC based alterations of the significantly different species in the **A**) gut microbiome between CMA and placebo groups on Day 70 vs Day 0; **B**) gut microbiome between Day 70 vs Day 0 in the CMA and placebo groups; **C**) oral microbiome between CMA and placebo groups on Day 70 vs Day 0; **D**) oral microbiome between Day 70 vs Day 0 in the CMA and placebo groups. Each dot represents a species and it has been colored according to its corresponding phylum. Asterisks indicate statistical significance based on paired Wilcoxon signed-rank test. p< 0.05. Log2FC: log2(fold change). **E**) Heatmaps show the correlations between the plasma levels of metabolites (other than lipids) and the abundance of the species in the gut microbiome. Asterisks indicate statistical significance based on Spearman correlation analysis. p < 0.05; Cor.Coeff: Correlation coefficient

Next, we compared the abundances of the species between CMA vs placebo groups on Day 70 in the gut microbiome. We found that the abundance of the butyrate-producing species (*Lawsonibacter asaccharolyticus, Eubacterium remulus* and *Eubacterium siraeum*) were significantly increased in the CMA group (Figure 5B, Dataset S16 & S17). Butyrate is a preferred energy source for gut epithelial cells, and it plays a vital role in maintaining health in humans. Notably, the abundance of acetaldehyde producer *Rothia mucilaginosa* was significantly downregulated in the gut microbiome of CMA group on Day 70 vs Day 0 (Figure 5A) and the placebo group on Day 70 vs Day 0 (Figure 5B, Dataset S16 & S17).

Similarly, we compared the differences in the species’ abundances between Day 70 vs Day 0 in the CMA and placebo groups in the oral microbiome. We found that the abundance of the specific species of Proteobacteria (*Cardiobacterium hominis*), Bacteriodetes (*Prevotella maculosa*) and Actinobacteria (*Corynebacterium matruchotii* and *Actinobaculum* sp oral taxon 183) were significantly decreased in the CMA group (Figure 5C & Figure S2, Dataset S18). We also found increased abundance of *Actinomyces* sp oral taxon 181 and reduced abundance of *Neisseria sicca* between CMA vs placebo groups on Day 70 in the oral microbiome (Figure 5D, Dataset S18).

We evaluated the association of the plasma levels of metabolites directly related with CMA with the abundance of species in the gut microbiome (Figure 5E) and oral microbiome (Figure S2) and found that *Faecalibacterium prausnitzii* is positively correlated with CMA related metabolites; namely cysteine, cysteinyl glycine, sarcosine, and N1-methylinosine (Figure 5E, Dataset S17). We also found that plasma cysteine levels are significantly positively correlated with the abundance of species associated with Firmicutes (*Roseburia faecis* and *Oscillibacter* sp 57 20) and Bacteriodetes (*Bacteroides ovatus* and *Bacteroides fragilis*). Additionally, we observed that the plasma levels of N1-methyl-4-pyridone-3-carboxamide and N1-methyl-2-pyridone-5-carboxamide are significantly positively correlated with the abundance of *Alistipes shahii* and negatively correlated with the abundance of *Bacteroides cellulosilyticus* and *Fusicatenibacter saccharivorans* (Figure 5E, Dataset S17).

Changes in the gut microbiome are linked to inflammation by triggering molecules involving interleukins and other cytokines(Schirmer *et al*, 2016). An increased abundance of *Coprococcus eutactus* and decreased abundance of *Bacteriodes faecis, Bacteriodes dorei, Bacteriodes xylanisolnes* and *Barnessiella intestihominis* in the gut microbiome were associated with FGF-21 levels (Figure 4D, Dataset S17). In the oral microbiome, we found that the abundances of *Prevotella jejuni* and *Alloprevotella tannerae* were significantly negatively correlated with the plasma level of FGF21 (Figure 4E, Dataset S18). Moreover, we found that increased abundance of *Prevotella copri* and *Neisseria sicca* and decreased abundance of *Neisseria elongate* and *Haemophilus parainfluenzae* in the oral microbiome associated with the plasma level of CCL23, of which gene expression level was significantly upregulated in NASH patients(Hart *et al*, 2017) (Figure 4E, Dataset S18). Interestingly, increased abundances of *Faecalibacterium prausnitzii* and *Bacteriodes fragilis* in the gut microbiome were positively correlated with the plasma level of CD8A, indicating the key role of the microbiome in the modulation of CD8 T cell responses (Figure 4D, S17).

Even though the oral and gut microbiome are distinct, a body of evidence claimed a strong relationship between these microenvironments due to the transfer of oral members to gut by a constant flow of saliva and ingestion(Schmidt *et al*, 2019). Therefore, we evaluated the species’ correlation in feces and saliva microbiome and revealed a significant positive correlation between species pairs, where only two pairs were negatively correlated (Figure S3). Interestingly, we found that the abundance of the species belongs to the Prevotella genus in the oral microbiome and the species belong to Bacteroides genus in the gut microbiome are affected apparently from each other’s abundance (Figure S3).

### The associations between clinical variables and multi-omics data

To study the link between improved clinical parameters and metabolism, we determined the significant correlations between improved clinical variables (e.g., hepatic fat, ALT, AST, and uric acid) and various metabolites (Figure 6A, Dataset S13). The plasma level of cysteine-glutathione disulfide, iminodiacetate, 4-hydroxychlorothalonil and arachidonoylcholine significantly and negatively correlated with hepatic fat (Figure 6A, Dataset S13). Of these metabolites, cysteine-glutathione disulfide significantly and negatively correlated with ALT, AST, and uric acid levels, indicating that glutathione metabolism has a key role in improving liver functions in NAFLD patients.

**Figure 6.**
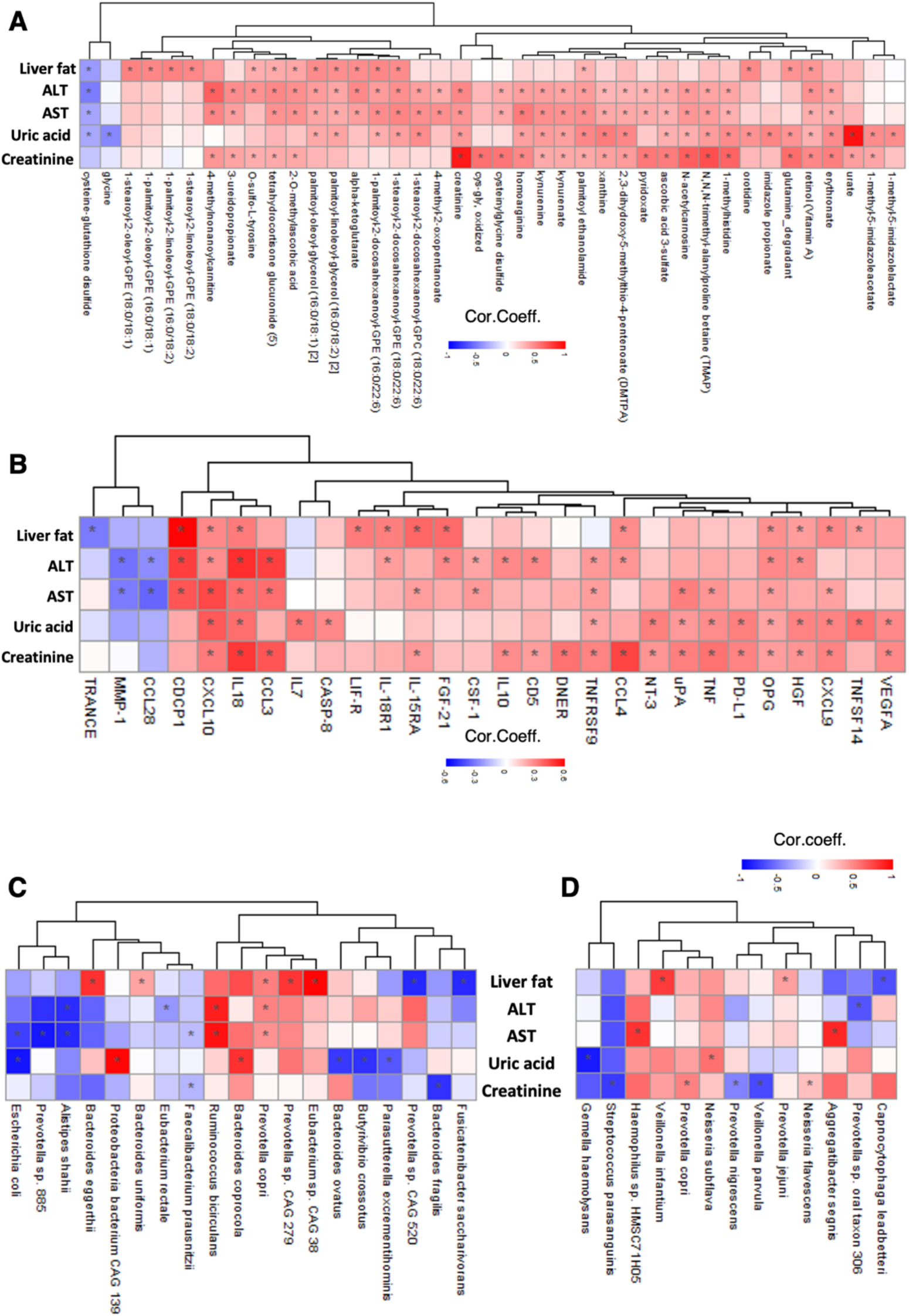
Associations between the Clinical Data and omics data. Heatmap shows the association between the plasma level of clinical variables including liver fat, ALT, AST, uric acid, and creatinine with **A)** plasma level of 10 most significant metabolites, **B)** plasma level of 10 most significant inflammation related proteins, **C)** the abundance of the species in gut microbiome and **D)** the abundance of the species in oral microbiome. AST, Aspartate aminotransferase; ALT, Alanine transaminase. Asterisks indicate statistical significance based on Spearman correlation analysis. p < 0.05; Cor.Coeff: Correlation coefficient.

We assessed the associations between improved clinical parameters and inflammation by identifying the significant correlations between improved clinical variables (e.g., hepatic fat, ALT, AST, and uric acid) and various inflammation-related proteins (Figure 6B, Dataset S14). We observed a positive correlation between the liver fat and the plasma level of CDCP1, CCL23, TNFSF14 and FGF-21; the AST level and the plasma level of CSF-1 and CDCP1; the ALT level and the plasma levels CDCP1, CSF-1 and FGF-21; the uric acid level and plasma level of TNFSF14 as well as the creatinine level and plasma levels of OSM and EN-RAGE. Additionally, we found a negative correlation between the levels of ALT and AST and plasma level of CCL28, which is significantly positively correlated with the plasma level of serine.

Correlation analysis between the improved clinical variables and the abundances of key discriminatory microbial species showed that the abundances of *Prevotella copri* in gut microbiome were positively correlated with liver fat, AST, ALT and systolic blood pressure level (Figure 6C, S17). Additionally, the abundance of *Prevotella* sp CAG 279, *Eubacterium* sp CAG 38, *Bacteroides uniformis* and *Bacteroides eggerthii* are other positively correlated species with the liver fat. In contrast, the abundance of *Fusicatenibacter saccharivorans* and *Prevotella* sp CAG 279 are negatively correlated with liver fat (Figure 6C, Dataset S17). We also found that the abundance of *Ruminoccocus bicirculans* was significantly correlated with ALT and AST levels, however, the abundance of *Alistipses shahii* showed a negative correlation with both ALT and AST levels (Figure 6C, Dataset S17). Of note, we found significant negative correlation *Eubacterium rectale* with ALT levels and *Faecalibacterium prausnitzii* with AST levels; both are well-known species associated with healthy gut microbiome (Figure 6C, Dataset S16 & S17).

Several studies reported that the oral microbiota reflects changes in gut microbiota’s dysbiosis, and it might be indicator for liver pathologies(Bajaj *et al*, 2015). In our study, the evaluation of oral microbiome showed that the abundances of *Veilonella infantum* and *Provetolla jejuni* are positively and the abundances of *Capnocytophaga leadbetteri* are negatively correlated with liver fat; AST levels were positively correlated with the abundances of *Aggretibacter segnis* and *Haemophilus* sp HMCS7 1H05; ALT levels were negatively correlated with the abundances of *Prevotella* oral taxon 306 (Figure 6D, Dataset S18).

It has been shown that gut microbiota plays significant roles in uric acid metabolism. Here, we found that the abundances of *Bacteroides coprocola* and *Proteobacteria bacterium* CAG 139 in the gut microbiome and *Neisseria subflava* in the oral microbiome were significantly positively correlated with the uric acid levels. In contrast, the abundances of *Bacteroides ovatus, Butyrivibrio crossotus* and *Parasutterella excremntihominis* in the gut microbiome and the abundances of *Gemella haemolysans* in the oral microbiome were significantly negatively correlated (Figure 6C and D, Dataset S16, S17 and S18). We also found that the abundances of the *Fusobacterium periodonticum* and *Porphyromonas somerae*, opportunistic pathogens in the oral microbiome were significantly positively correlated to the urea levels (Figure S2A). Interestingly, creatinine levels correlated with increased abundances of *Prevotella copri* and *Neisseria flavescens* and decreased abundances of *Prevotella nigrescens* and *Veilonella parvula* in the oral microbiome (Figure 6D, Dataset S18). The abundances of *Bacteroides fragilis* and *Faecalibacterium prausnitzii* in the gut microbiome are negatively correlated with creatinine levels (Figure 6C, Dataset S17).

### Integrative Multi-Omics Analysis

We generated an integrative multi-omics network based on clinical variables, proteomics, metabolomics and microbiome data to show the functional relationship between analytes within and between omics data. The filtered network (edges FDR < 0.05) can be viewed at iNetModels (http://inetmodels.com), an open-access interactive web platform for multi-omics data visualization and database. We overlapped the statistically altered metabolites, proteins and clinical variables to the sub-network of the metabolic activators and liver fat, as primary clinical variable targeted by CMA (Figure 7A). Furthermore, we performed a centrality analysis of the filtered network to identify the most central analytes. We identified carnitine as the most prominent metabolic activator. We also observed that top 20-degree metabolites were dominated by lipid structures (sphingomyelin, phosphatidylinositol/choline, diacylglycerol, and fatty acids), gamma-glutamylisoleucine, and retinol (Vitamin A). For the clinical variables, the level of uric acid, that was shown to be downregulated in the CMA group, was identified as one of the top 5 most connected clinical nodes.

**Figure 7.**
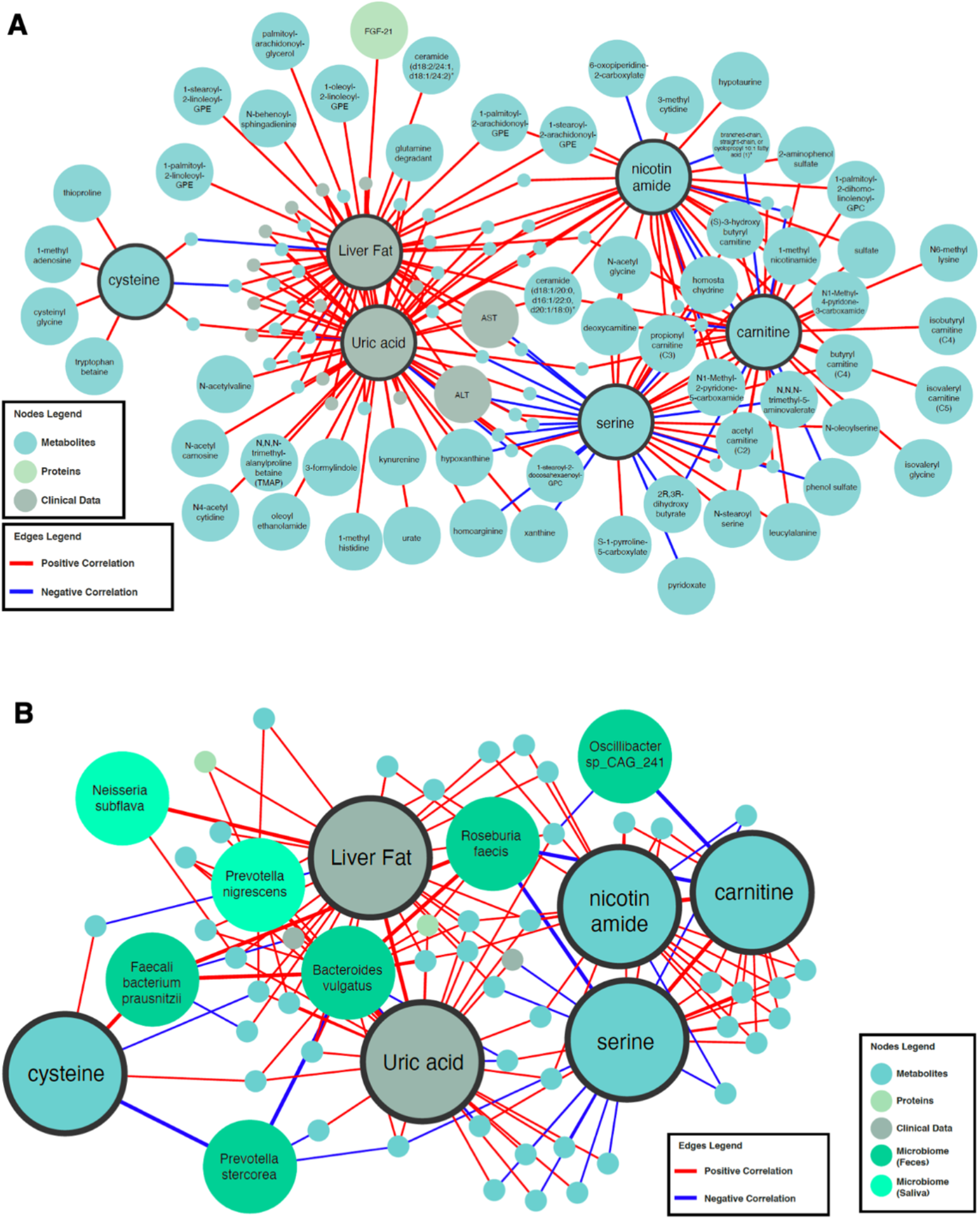
Integrating omics data based on Network analysis. **A)** Neighbors of the CMA, including serine, carnitine, nicotinamide and cysteine, and two important clinical variables (Liver Fat and Uric Acid) are presented based on the multi-omics network analysis. Only analytes that are significantly altered in CMA Day 70 vs Day 0 are highlighted. **B)** First degree of microbial neighbors of the CMA and two important clinical variables (Liver Fat and Uric Acid) are presented based the multi-omics network analysis. Full networks can be found in iNetModels (http://inetmodels.com).

Moreover, we observed that the abundance of the *Roseburia faecis* (significantly downregulated in the CMA group) is significantly negatively correlated with the plasma level of serine and carnitine (Figure 7B). Based on these results, the multi-omics integrative network analysis complemented and strengthened the single-omics analysis findings. We also observed that serine and carnitine play key roles among the metabolic activators in improving the clinical outcome.

### Safety

Both the CMA and placebo treatments were generally well tolerated. Eighteen patients (12 in the CMA group and 6 in the placebo group) reported only mild-to-moderate adverse events, including nonspecific gastrointestinal (33%), respiratory (23%), and musculoskeletal (20%) symptoms, and all patients decided to complete the study (Dataset S15). Gastrointestinal symptoms in three patients in the placebo group and one in the CMA group were associated with the intervention, and their drug doses were reduced to one per day. Headache, rash, gout monoarthropathy, eye disorders, and erectile dysfunction were reported in the CMA group, and the renal stone was reported in the placebo group but did not appear to be related to the treatment.

## DISCUSSION

Based on data-driven modelling and systems biology, we have identified a CMA based on four components to increase the liver’s fat oxidation and enhance the mitochondrial function in the cells, thus activating hepatic mitochondrial fatty acid uptake and β-oxidation along with reducing oxidative stress. To test this hypothesis in a clinical setting, we performed on a randomized, placebo-controlled phase 2 study to investigate the efficacy and safety of CMA in NAFLD patients. In this study, we found that CMA (10%) reduces the hepatic fat content assessed by MRI-PDFF after 70 days, the primary outcome variable. Secondary outcomes were also significantly improved, including reductions in serum ALT (39%) and AST (30%) levels. Also reduced were CD8A, CSF-1, CCL23, and OSM levels, as shown by plasma proteomics analysis, indicating that CMA attenuates hepatic inflammation. The positive results were not linked to significant weight loss in the CMA group.

Analysis of untargeted metabolomics data confirmed the expected biological outcomes of CMA treatment. Levels of plasma N1-methyl-2-pyridone-5-carboxamide, and 1-methylnicotinamide were increased, suggesting that nicotinamide riboside provides sufficient substrate and NAD^+^ for mitochondrial fatty acid oxidation. These metabolites have shown to be increased in a recent study, supporting the insulin-sensitizing ability of the nicotinamide mononucleotide, another NAD+ intermediate, in skeletal muscle of prediabetic women (Yoshino *et al*, 2021). Plasma levels of N-acetylglycine, N-palmitoylserine, N-oleoylserine, and N-stearoylserine were also increased, suggesting that CMA treatment improves the serine deficiency associated with hepatic steatosis. More importantly, fatty acid oxidation and carnitine metabolism were remarkably facilitated, as judged from the robust plasma levels of deoxycarnitine, acetylcarnitine, and butyrylcarnitine. Moreover, after weight loss adjustment, the levels of tryptophan metabolites, including kynurenate, kynurenine, and tryptophan betaine, decreased significantly after CMA treatment.

Unexpectedly, CMA rapidly reduced uric acid levels. Significantly decreased metabolites associated with the uric acids were shown (Figure S4). Uric acid stimulates hepatic steatosis either directly or by activating NLRP3 inflammasomes (Lanaspa *et al*, 2012; Wan *et al*, 2016). Although the extent to which uric acid reduction contributed to the regression in hepatic steatosis is unclear, it likely had at least an additive effect. How might CMA reduce serum uric acid levels in overweight and obese subjects? A recent study revealed a strong association with uric acid and 1-methyhistidine levels, which is significantly decreased with CMA therapy (Pietzner *et al*, 2021). Another possible explanation is that the glycine derived from serine, lowers uric acid levels by stimulating uricosuria (Oshima *et al*, 2019). Given the significantly reduced level of purine metabolism, it is also possible that L-carnitine inhibited xanthine oxidase activity and uric acid production (Volek *et al*, 2002). Further clinical studies that include hyperuricemic patients will be required to determine whether this effect can be generalized to treat patients with hyperuricemia. The direct or indirect impact of the microbiome on NAFLD remains poorly understood. In this study, we comprehensively characterized the gut and oral microbiome in NAFLD patients before and after administration of metabolic activators. Our results collectively indicated that CMA induced literature supported beneficial alterations such as positive correlation with the abundances of *Faecalibacterium prausnitzii* (Grabherr *et al*, 2019; Munukka *et al*, 2017), *Roseburia faecis (Tamanai-Shacoori et al, 2017)* or butyrate producers (Baxter *et al*, 2019; Boesmans *et al*, 2018). In addition to identifying the key species, defining the distinct signatures of the microbiome and liver-related clinical parameters in the well-defined patient cohort is valuable in searching for new biomarkers for NAFLD.

A few limitations of the study need to be considered. First, the study was designed to include 45 participants; however, because of COVID-19 restrictions, only 32 subjects were enrolled. Second, patients who had type 2 diabetes or dyslipidemia or were taking anti-diabetic medications were excluded, which resulted in a more homogenous clinical trial group. Third, hepatic fat content and inflammation were assessed by MRI-based methods and plasma inflammatory protein markers rather than liver histology. Our findings warrant a clinical trial in patients with biopsy-proven nonalcoholic steatohepatitis to delineate the effects CMA on hepatic injury and inflammation.

The safety profile of CMA in these patients was consistent with the results of our previous one-day calibration study and clinical trials, including only a single component of the CMA (Zhang *et al*., 2020). Our present study showed that CMA was safe and well-tolerated in patients with NAFLD, and no major safety concerns were identified. In conclusion, CMA significantly reduced hepatic fat content and serum markers of hepatic injury in 10 weeks. These findings suggest that targeting multiple pathways by CMA is a potentially effective therapeutic strategy for NAFLD.

## METHODS

### Trial Design and Oversight

Patients for this randomized, placebo-controlled, phase 2 study were recruited at the Koç University Hospital, Istanbul, Turkey (Dataset S1). The trial was conducted following Good Clinical Practice guidelines and the principles of the Declaration of Helsinki. An independent external data monitoring committee oversaw the safety of the participants and the risk-benefit analysis. Written informed consent was obtained from all participants before trial-related procedures were initiated. The study was approved by the ethics committee at Koç University (Decision No: 2018.351.IRB1.043). The study was registered at https://eudract.ema.europa.eu/ with the accession number EudraCT_2018-000894-59 and at https://clinicaltrials.gov/ with the accession number NCT04330326.

### Participants

Overweight or obese patients 18–70 years of age were enrolled in the trial if they were diagnosed with NAFLD and met all the inclusion criteria: body mass index >27 kg/m^2^, triglycerides ≤354 mg/dl, low-density lipoprotein cholesterol ≤175 mg/dl, and increased hepatic fat (>5.5%). Patients were excluded if they carried the PNPLA3 I148M (homozygous for I148M), had ALT or AST levels >3-fold higher than the upper limit of normal, or had taken oral antidiabetics, including metformin, within three months. The inclusion and exclusion criteria are detailed in the Supplementary Appendix. The main characteristics of the study participants are presented in Table 1 and Dataset S2.

### Randomization, Interventions, and Follow-up

Patients were randomly assigned to CMA or placebo (2:1). Patient information (patient number, date of birth, initials) was entered into the web-based randomization system, and the randomization codes were entered into the electronic case report form.

CMA treatment was given for 70 days after the initial diagnosis of high hepatic fat by MRI-PDFF. Patients in the treatment group took one dose of CMA (3.73 g L-carnitine tartrate, 1 g nicotinamide riboside, 12.35 g serine, and 2.55 g N-acetyl-l-cysteine) daily for the first 14 days (after dinner) and two doses daily for the next 56 days (after breakfast and dinner). Further information is provided in the Supplementary Appendix.

The subjects returned to the study centre for complete follow-up evaluations, including body composition analysis and adverse events recording. Hepatic fat was determined by MRI-PDFF on Days 0, 14 and 70. Plasma samples for proteomics and metabolomics analyses were obtained on Days 0, 14, and 70. After Day 70, subjects stopped taking their drugs (Dataset S1).

### Study Outcomes

The study’s primary objective was to assess the difference in hepatic fat content, quantified by MRI-PDFF, between subjects treated for 70 days with CMA or placebo (Dataset S3). The secondary objectives were to assess the tolerability and safety profile of CMA, as judged by laboratory analyses and physical parameters (Dataset S4), and to examine additional efficacy parameters, as evaluated by biochemical, metabolomic and proteomics analysis.

The number and characteristics of adverse events, serious adverse events, and treatment discontinuation due to study drugs were reported from the beginning of the study to the end of the follow-up period as key safety endpoints (Supplementary Appendix). Changes in vital signs (systolic and diastolic blood pressures, pulse, respiratory rate, body temperature, pulse oximetry values), baseline values, and treatment status were recorded by phone contacts between the visits. A complete list of endpoints is provided in the Supplementary Appendix.

### Untargeted Metabolomics Analysis

Plasma samples were collected on Days 0, 14, and 70 (Datasets S5–S8) for nontargeted metabolite profiling by Metabolon (Durham, NC). The samples were prepared with an automated system (MicroLab STAR, Hamilton Company, Reno, NV). For quality control purposes, a recovery standard was added before the first step of the extraction. To remove protein and dissociated small molecules bound to protein or trapped in the precipitated protein matrix, and to recover chemically diverse metabolites, proteins were precipitated with methanol under vigorous shaking for 2 min (Glen Mills GenoGrinder 2000) and centrifuged. The resulting extract was divided into four fractions: one each for analysis by ultraperformance liquid chromatography-tandem mass spectroscopy (UPLC-MS/MS) with positive ion-mode electrospray ionization, UPLC-MS/MS with negative ion-mode electrospray ionization, and gas chromatography-mass spectrometry; one fraction was reserved as a backup.

### Inflammatory Protein Markers

In the plasma samples, inflammatory protein markers were determined with the Olink Inflammation panel (Olink Bioscience, Uppsala, Sweden) (Datasets S9–S11). Briefly, each sample was incubated with 92 DNA-labeled antibody pairs (proximity probes). When an antibody pair binds to its corresponding antigens, the corresponding DNA tails form an amplicon by proximity extension, which can be quantified by high-throughput, real-time PCR. Probe solution (3 μl) was mixed with 1 μl of sample and incubated overnight at 4°C. Then 96 μl of extension solution containing extension enzyme and PCR reagents for the pre-amplification step was added. The extension products were mixed with detection reagents and primers and loaded on the chip for qPCR analysis with the BioMark HD System (Fluidigm Corporation, USA).To minimize inter and intra-run variation, the data were normalized to both an internal control and an interpolate control. Normalized data were expressed in arbitrary units (Normalized Protein eXpression, NPX) on a log2 scale and linearized with the formula 2^NPX^. A high NPX indicates a high protein concentration. The limit of detection, determined for each of the 92 assays, was defined as three standard deviations above the negative control (background).

### Statistical Analysis

Values are expressed as the mean ± standard deviation (SD) (continuous variables) or as n (%). Safety and exploratory efficacy endpoints were analyzed in all randomized patients who received CMA. Differences in clinical and physiological variables between time points were analyzed by paired *t-test*, followed by one-way ANOVA to find shifts between CMA and placebo groups at each time point. Missing values were removed in a pairwise fashion.

For analysis of plasma metabolomics, raw data from Day 14 and Day 70 in both groups were scaled to data on Day 0. Metabolite changes in the CMA group vs the placebo group over time (Dataset S6) were analyzed by one-way ANOVA. Differences between the CMA and placebo groups on Day 0, Day 14 and Day 70 were also calculated (Dataset S7). Metabolites with a false-discovery rate of 5% were considered statistically significant. The calculation was adjusted for weight loss in a linear model using the limma package in R (v4.0.2). Differences were considered significant at p < 0.05.

For analysis of plasma proteomics, raw data from Day 14 and Day 70 were scaled to data on Day 0. Subsequently, ANOVA was used to calculate the CMA group changes vs placebo over time (Dataset S10). P < 0.05 was considered statistically significant.

Finally, Spearman correlation analysis (false discovery rate <0.05) was used to analyze the association between individual components of CMA and other metabolites (Dataset S8) and between the significantly altered clinical parameters (e.g., hepatic fat and plasma levels of ALT, AST, uric acid, and creatinine) and other metabolites (Dataset S13). Spearman correlation analysis was also used to study the associations between the plasma protein levels and the plasma metabolite levels (Datasets S12).

### Metagenomics Data Analysis

Fresh stool and saliva specimens were collected and preserved using DNA/RNA Shield Fecal Collection tubes (Zymo Research, Irvine, CA) and DNA/RNA Shield Collection Tube (Zymo Research, Irvine, CA), respectively. DNA extractions from the faecal samples were done using QIAamp PowerFecal Pro DNA Kit (Qiagen, Hilden, Germany) and the saliva samples using QIAamp DNA Microbiome Kit (Qiagen, Hilden, Germany). All protocol procedures were performed according to the manufacturer’s instructions. Quantification of extracted DNA was determined fluorometrically on the Qubit® 3.0 Fluorometer (Thermo Fisher Scientific, United States) using the QubitTM dsDNA HS Assay Kit. DNA purity was determined via 260/280 and 260/230 ratios measured on the NanoDrop 1000 (Thermo Fisher Scientific, United States). The SMARTer Thruplex DNA-Seq (Takara Bio) was used for library preparation (Option:350 bp; Category: low input). Samples were sequenced on NovaSeq6000(NovaSeq Control Software 1.7.0/RTA v3.4.4) with a 151nt (Read1)-10nt(Index1)-10nt(Index2)-151nt(Read2) setup using ‘NovaSeqXp’ workflow in ‘S4’ mode flow cell. The Bcl to FastQ conversion was performed using bcl2fastq_v2.20.0.422 from the CASAVA software suite. The quality scale used is Sanger /phred33/Illumina 1.8+.

Raw paired-end metagenomics data were analysed using Metaphlan3 (Beghini *et al*, 2020) to extract each sample’s taxonomic profiles. The abundant data were then analysed using the Wilcoxon rank-sum test to identify the species different between subjects with no steatosis compared to the other groups. Spearman correlation analysis was used to analyse the associations between selected analytes and the taxonomic abundance data. The correlation between oral and gut metagenomics data (by filtering the species with abundance > 1% in at least 5 data points). The *SciPy* package was used. All analyses were done using Python 3.

### Generation of Multi-Omics Network

Multi-omics network was generated based on the Spearman correlations, and the significant associations (FDR < 0.05) are presented. The analyses were performed with the SciPy package in Python 3.7. Centrality analysis on the network was performed using iGraph Python.

## Data Availability

All data associated with this study are available in the main text or the supplementary materials.

## Author Contributions

M.U., J.B., and A.M. designed the study; M.Z., M.A., B.S., and M.G.G. recruited patients; H.Y., O.A., M.A., C.F., W.K., X.L., J.M.S., C.Z., S.S., and J.N. did the experiments; H.Y., O.A., M.A., C.F., W.K., X.L., J.M.S., C.Z., S.S., J.N., and A.M analyzed the data; M.Z., M.U., J.B., and A.M. interpreted the data; M.Z, O.A., and A.M. wrote the manuscript; and all authors helped revise the manuscript.

## Data and materials availability

All data associated with this study are available in the main text or the supplementary materials.

## REFERENCES

Abeysekera KWM, Fernandes GS, Hammerton G, Portal AJ, Gordon FH, Heron J, Hickman M (2020) Prevalence of steatosis and fibrosis in young adults in the UK: a population-based study. The Lancet Gastroenterology & Hepatology 5: 295–305

Alferink LJ, Kiefte-de Jong JC, Erler NS, Veldt BJ, Schoufour JD, de Knegt RJ, Ikram MA, Metselaar HJ, Janssen H, Franco OH et al (2019) Association of dietary macronutrient composition and non-alcoholic fatty liver disease in an ageing population: the Rotterdam Study. Gut 68: 1088–1098

Anstee QM, Reeves HL, Kotsiliti E, Govaere O, Heikenwalder M (2019) From NASH to HCC: current concepts and future challenges. Nature Reviews Gastroenterology & Hepatology 16: 411–428

Aron-Wisnewsky J, Vigliotti C, Witjes J, Le P, Holleboom AG, Verheij J, Nieuwdorp M, Clément K (2020) Gut microbiota and human NAFLD: disentangling microbial signatures from metabolic disorders. Nature Reviews Gastroenterology & Hepatology 17: 279–297

Bajaj JS, Betrapally NS, Hylemon PB, Heuman DM, Daita K, White MB, Unser A, Thacker LR, Sanyal AJ, Kang DJ et al (2015) Salivary microbiota reflects changes in gut microbiota in cirrhosis with hepatic encephalopathy. Hepatology 62: 1260–1271

Barbier-Torres L, Fortner KA, Iruzubieta P, Delgado TC, Giddings E, Chen Y, Champagne D, Fernández-Ramos D, Mestre D, Gomez-Santos B et al (2020) Silencing hepatic MCJ attenuates non-alcoholic fatty liver disease (NAFLD) by increasing mitochondrial fatty acid oxidation. Nature Communications 11: 3360

Barbier-Torres L, Iruzubieta P, Fernández-Ramos D, Delgado TC, Taibo D, Guitiérrez-de-Juan V, Varela-Rey M, Azkargorta M, Navasa N, Fernández-Tussy P et al (2017) The mitochondrial negative regulator MCJ is a therapeutic target for acetaminophen-induced liver injury. Nature Communications 8: 2068

Baxter NT, Schmidt AW, Venkataraman A, Kim KS, Waldron C, Schmidt TM (2019) Dynamics of Human Gut Microbiota and Short-Chain Fatty Acids in Response to Dietary Interventions with Three Fermentable Fibers. mBio 10: e02566–02518

Beghini F, McIver LJ, Blanco-Míguez A, Dubois L, Asnicar F, Maharjan S, Mailyan A, Thomas AM, Manghi P, Valles-Colomer M et al (2020) Integrating taxonomic, functional, and strain-level profiling of diverse microbial communities with bioBakery 3. bioRxiv: 2020.2011.2019.388223

Boesmans L, Valles-Colomer M, Wang J, Eeckhaut V, Falony G, Ducatelle R, Van Immerseel F, Raes J, Verbeke K (2018) Butyrate Producers as Potential Next-Generation Probiotics: Safety Assessment of the Administration of Butyricicoccus pullicaecorum to Healthy Volunteers. mSystems 3: e00094–00018

Bugianesi E (2020) Fatty liver disease: putting the spotlight on a silent menace for young adults. The Lancet Gastroenterology & Hepatology 5: 236–238

Dushay J, Chui PC, Gopalakrishnan GS, Varela-Rey M, Crawley M, Fisher FM, Badman MK, Martinez-Chantar ML, Maratos-Flier E (2010) Increased fibroblast growth factor 21 in obesity and nonalcoholic fatty liver disease. Gastroenterology 139: 456–463

Ertunc ME, Hotamisligil GS (2016) Lipid signaling and lipotoxicity in metaflammation: indications for metabolic disease pathogenesis and treatment. J Lipid Res 57: 2099–2114

Estes C, Razavi H, Loomba R, Younossi Z, Sanyal AJ (2018) Modeling the epidemic of nonalcoholic fatty liver disease demonstrates an exponential increase in burden of disease. Hepatology 67: 123–133

Friedman SL, Neuschwander-Tetri BA, Rinella M, Sanyal AJ (2018) Mechanisms of NAFLD development and therapeutic strategies. Nat Med 24: 908–922

Grabherr F, Grander C, Effenberger M, Adolph TE, Tilg H (2019) Gut Dysfunction and Non-alcoholic Fatty Liver Disease. Front Endocrinol (Lausanne) 10: 611

Guohong L, Qingxi Z, Hongyun W (2019) Characteristics of intestinal bacteria with fatty liver diseases and cirrhosis. Annals of Hepatology 18: 796–803

Hart KM, Fabre T, Sciurba JC, Gieseck RL, Borthwick LA, Vannella KM, Acciani TH, de Queiroz Prado R, Thompson RW, White S et al (2017) Type 2 immunity is protective in metabolic disease but exacerbates NAFLD collaboratively with TGF-β. Science Translational Medicine 9: eaal3694

Haukka JK, Sandholm N, Forsblom C, Cobb JE, Groop P-H, Ferrannini E (2018) Metabolomic Profile Predicts Development of Microalbuminuria in Individuals with Type 1 Diabetes. Scientific Reports 8: 13853

Lanaspa MA, Sanchez-Lozada LG, Choi YJ, Cicerchi C, Kanbay M, Roncal-Jimenez CA, Ishimoto T, Li N, Marek G, Duranay M et al (2012) Uric acid induces hepatic steatosis by generation of mitochondrial oxidative stress: potential role in fructose-dependent and -independent fatty liver. J Biol Chem 287: 40732–40744

Lee S, Zhang C, Kilicarslan M, Piening Brian D, Bjornson E, Hallström Björn M, Groen Albert K, Ferrannini E, Laakso M, Snyder M et al (2016) Integrated Network Analysis Reveals an Association between Plasma Mannose Levels and Insulin Resistance. Cell Metabolism 24: 172–184

Lovric A, Granér M, Bjornson E, Arif M, Benfeitas R, Nyman K, Ståhlman M, Pentikäinen MO, Lundbom J, Hakkarainen A et al (2018) Characterization of different fat depots in NAFLD using inflammation-associated proteome, lipidome and metabolome. Scientific Reports 8: 14200

Mardinoglu A, Agren R, Kampf C, Asplund A, Uhlen M, Nielsen J (2014) Genome-scale metabolic modelling of hepatocytes reveals serine deficiency in patients with non-alcoholic fatty liver disease. Nature Communications 5: 3083

Mardinoglu A, Bjornson E, Zhang C, Klevstig M, Söderlund S, Ståhlman M, Adiels M, Hakkarainen A, Lundbom N, Kilicarslan M et al (2017) Personal model-assisted identification of NAD(+) and glutathione metabolism as intervention target in NAFLD. Mol Syst Biol 13: 916

Mardinoglu A, Boren J, Smith U, Uhlen M, Nielsen J (2018a) Systems biology in hepatology: approaches and applications. Nature Reviews Gastroenterology & Hepatology 15: 365–377

Mardinoglu A, Ural D, Zeybel M, Yuksel HH, Uhlén M, Borén J (2019) The Potential Use of Metabolic Cofactors in Treatment of NAFLD. Nutrients 11: 1578

Mardinoglu A, Wu H, Bjornson E, Zhang C, Hakkarainen A, Räsänen SM, Lee S, Mancina RM, Bergentall M, Pietiläinen KH et al (2018b) An Integrated Understanding of the Rapid Metabolic Benefits of a Carbohydrate-Restricted Diet on Hepatic Steatosis in Humans. Cell Metab 27: 559-571.e555

Marra F, Lotersztajn S (2013) Pathophysiology of NASH: perspectives for a targeted treatment. Curr Pharm Des 19: 5250–5269

Munukka E, Rintala A, Toivonen R, Nylund M, Yang B, Takanen A, Hänninen A, Vuopio J, Huovinen P, Jalkanen S et al (2017) Faecalibacterium prausnitzii treatment improves hepatic health and reduces adipose tissue inflammation in high-fat fed mice. ISME J 11: 1667–1679

Oshima S, Shiiya S, Nakamura Y (2019) Serum Uric Acid-Lowering Effects of Combined Glycine and Tryptophan Treatments in Subjects with Mild Hyperuricemia: A Randomized, Double-Blind, Placebo-Controlled, Crossover Study. Nutrients 11

Pietzner M, Stewart ID, Raffler J, Khaw K-T, Michelotti GA, Kastenmüller G, Wareham NJ, Langenberg C (2021) Plasma metabolites to profile pathways in noncommunicable disease multimorbidity. Nature Medicine 27: 471–479

Porras D, Nistal E, Martínez-Flórez S, González-Gallego J, García-Mediavilla MV, Sánchez-Campos S (2018) Intestinal Microbiota Modulation in Obesity-Related Non-alcoholic Fatty Liver Disease. Frontiers in physiology 9: 1813–1813

Romero-Gómez M, Zelber-Sagi S, Trenell M (2017) Treatment of NAFLD with diet, physical activity and exercise. J Hepatol 67: 829–846

Rusli F, Deelen J, Andriyani E, Boekschoten MV, Lute C, van den Akker EB, Müller M, Beekman M, Steegenga WT (2016) Fibroblast growth factor 21 reflects liver fat accumulation and dysregulation of signalling pathways in the liver of C57BL/6J mice. Sci Rep 6: 30484

Sanyal AJ (2019) Past, present and future perspectives in nonalcoholic fatty liver disease. Nat Rev Gastroenterol Hepatol 16: 377–386

Schirmer M, Smeekens SP, Vlamakis H, Jaeger M, Oosting M, Franzosa EA, Ter Horst R, Jansen T, Jacobs L, Bonder MJ et al (2016) Linking the Human Gut Microbiome to Inflammatory Cytokine Production Capacity. Cell 167: 1125-1136.e1128

Schmidt TSB, Hayward MR, Coelho LP, Li SS, Costea PI, Voigt AY, Wirbel J, Maistrenko OM, Alves RJC, Bergsten E et al (2019) Extensive transmission of microbes along the gastrointestinal tract. Elife 8

Tamanai-Shacoori Z, Smida I, Bousarghin L, Loreal O, Meuric V, Fong SB, Bonnaure-Mallet M, Jolivet-Gougeon A (2017) Roseburia spp.: a marker of health? Future Microbiology 12: 157–170

Velenosi TJ, Thomson BKA, Tonial NC, RaoPeters AAE, Mio MA, Lajoie GA, Garg AX, House AA, Urquhart BL (2019) Untargeted metabolomics reveals N, N, N-trimethyl-L-alanyl-L-proline betaine (TMAP) as a novel biomarker of kidney function. Sci Rep 9: 6831

Volek JS, Kraemer WJ, Rubin MR, Gómez AL, Ratamess NA, Gaynor P (2002) l-Carnitine l-tartrate supplementation favorably affects markers of recovery from exercise stress. American Journal of Physiology-Endocrinology and Metabolism 282: E474–E482

Wan X, Xu C, Lin Y, Lu C, Li D, Sang J, He H, Liu X, Li Y, Yu C (2016) Uric acid regulates hepatic steatosis and insulin resistance through the NLRP3 inflammasome-dependent mechanism. Journal of Hepatology 64: 925–932

Yoshino M, Yoshino J, Kayser BD, Patti G, Franczyk MP, Mills KF, Sindelar M, Pietka T, Patterson BW, Imai SI et al (2021) Nicotinamide mononucleotide increases muscle insulin sensitivity in prediabetic women. Science

Younossi Z, Anstee QM, Marietti M, Hardy T, Henry L, Eslam M, George J, Bugianesi E (2018) Global burden of NAFLD and NASH: trends, predictions, risk factors and prevention. Nat Rev Gastroenterol Hepatol 15: 11–20

Zhang C, Bjornson E, Arif M, Tebani A, Lovric A, Benfeitas R, Ozcan M, Juszczak K, Kim W, Kim JT et al (2020) The acute effect of metabolic cofactor supplementation: a potential therapeutic strategy against non-alcoholic fatty liver disease. Mol Syst Biol 16: e9495

Zhao M, Zhao L, Xiong X, He Y, Huang W, Liu Z, Ji L, Pan B, Guo X, Wang L et al (2020) TMAVA, a Metabolite of Intestinal Microbes, Is Increased in Plasma From Patients With Liver Steatosis, Inhibits γ-Butyrobetaine Hydroxylase, and Exacerbates Fatty Liver in Mice. Gastroenterology 158: 2266-2281.e2227

