## Supplementary_ for "Combined Metabolic Activators Reduces Liver Fat in Nonalcoholic Fatty Liver Disease Patients": 210519 Zeybel et al_supplementaryDatasetsLegends_E.docx

**SUPPLEMENTARY DATASET LEGENDS**

**Dataset S1.** Collection of plasma samples for proteomics and metabolomics analysis and feces and saliva samples for metagenomics analysis before and after treatment.

**Dataset S2.** Patient characteristics before and after treatment.

**Dataset S3.** Liver fat content of each patient before and after treatment.

**Dataset S4.** Summary of laboratory and physical variables before and after treatment.

**Dataset S5.** Untargeted metabolomics data for each patient before and after treatment.

**Dataset S6.** Levels of plasma metabolites that differed significantly between CMA and placebo groups on Day 14 and Day 70 vs Day 0 before and after weight loss adjustment. Only metabolites detected in >50% of samples were analyzed.

**Dataset S7.** Plasma metabolites that differed significantly between CMA and placebo groups on Days 0, 14, and Day 70 both before and after weight loss adjustment. Only metabolites detected in >50% of samples were analyzed.

**Dataset S8.** The associations between the plasma level of all metabolites with the plasma levels of metabolic activators including serine, carnitine, nicotinamide, and cysteine.

**Dataset S9.** Plasma proteomics data generated with the Olink inflammation panel before and after treatment.

**Dataset S10.** Inflammation related plasma protein levels that differed between the CMA and placebo groups on Day 0, Day 14, and Day 70. Only proteins detected in >50% samples were analyzed.

**Dataset S11.** The associations between the plasma level of all Inflammation related proteins with the plasma levels of metabolic activators including serine, carnitine, nicotinamide, and cysteine.

**Dataset S12.** All plasma metabolites that correlated with significantly altered plasma proteins, including CD8A, CSF-1, CCL23, FGF-21, and OSM.

**Dataset S13** All plasma metabolites that correlated with significantly altered clinical variables, including liver fat content and plasma levels of ALT, AST, uric acid, and creatinine

**Dataset S14** All Inflammation related plasma proteins that correlated with significantly altered clinical variables, including liver fat content and plasma levels of ALT, AST, uric acid, and creatinine

**Dataset S15.** Mild-to-moderate adverse events reported throughout the study by NAFLD patients.

**Dataset S16.** The metadata and abundance of species for feces and saliva samples. Feces and saliva samples were collected for generation of metagenomics data. Metadata for each sample analyzed in the study as well as the abundances of the species in gut and oral microbiome is presented.

**Dataset S17.** Statistical analysis of metagenomics data from feces samples. **A)** The differences between the abundances of species in gut microbiome between Day 70 vs Day 0 in the CMA/placebo groups as well as between the CMA and placebo groups on Day 70 and Day 0 are presented. Statistical significance calculated based on paired Wilcoxon signed-rank test (for group comparisons) and Wilcoxon rank-sum test (for timepoints comparisons). The associations between the abundance of species in gut microbiome and **B)** the plasma level of significant metabolites, **C)** the plasma level of all inflammation related proteins and **D)** physical and clinical variables are presented.

**Dataset S18.** Statistical analysis of metagenomics data from saliva samples. **A)** The differences between the abundances of species in oral microbiome between Day 70 vs Day 0 in the CMA/placebo groups as well as between the CMA and placebo groups on Day 0/Day 70 are presented. Statistical significance calculated based on paired Wilcoxon signed-rank test (for group comparisons) and Wilcoxon rank-sum test (for timepoints comparisons). The associations between the abundance of species in oral microbiome and **B)** the plasma level of significant metabolites, **C)** the plasma level of all inflammation related proteins and **D)** physical and clinical variables are presented.
