## Supplementary_ for "Combined Metabolic Activators Reduces Liver Fat in Nonalcoholic Fatty Liver Disease Patients": 210519 Zeybel et al_supplementaryFigures_E.docx

**Figure S1.**

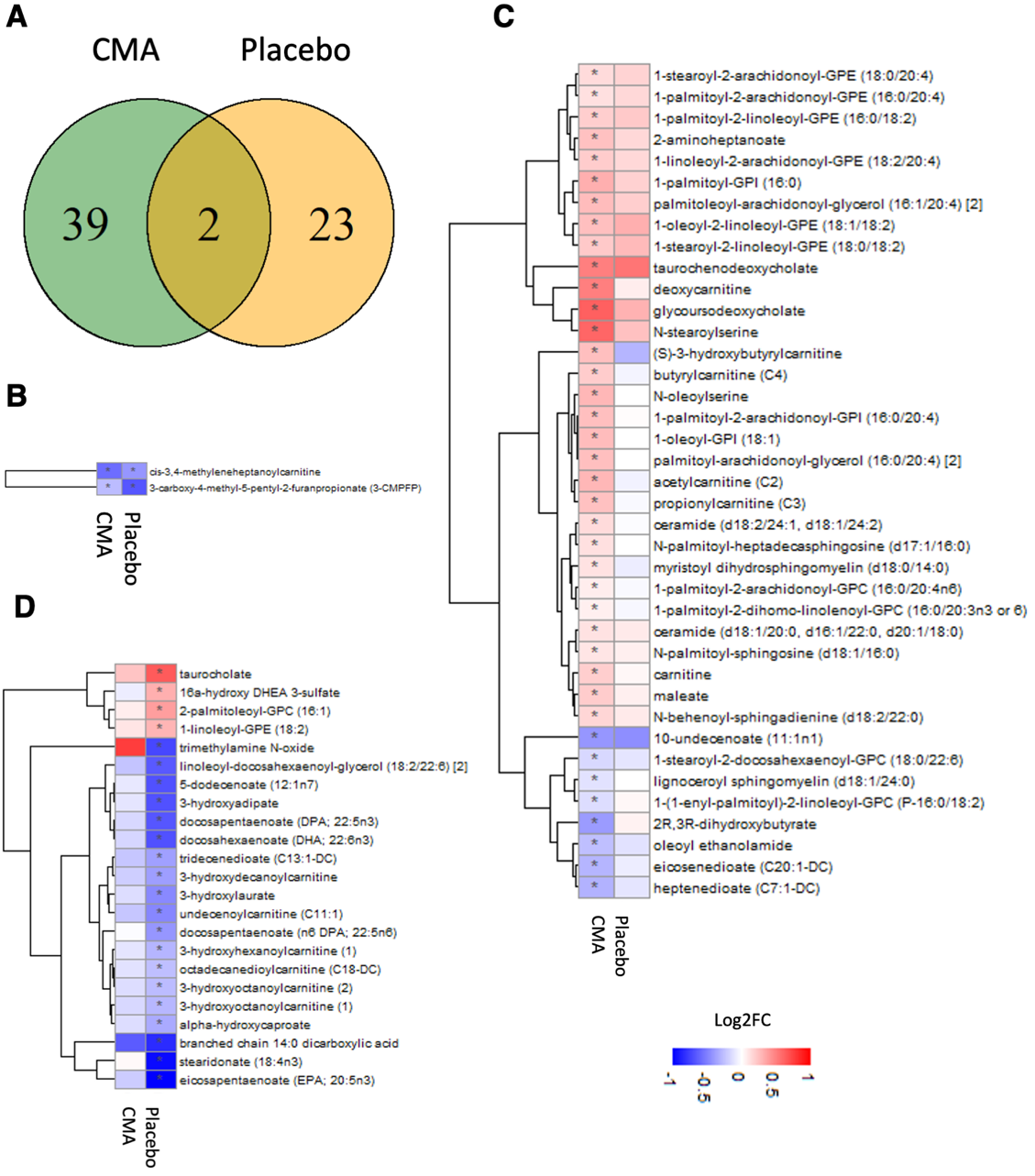

**Figure S1.** **CMA Effects the Plasma Level of Lipids**

Plasma level of lipids that are significantly different on Days 70 vs Day 0 in the CMA and placebo groups after weight loss adjustments. **A)** Venn-diagram representing the number of identified lipids that are significantly different on Days 70 vs Day 0 in the CMA and placebo groups. The intersection represents lipids that are significantly different in both groups. Association between the plasma level of significantly different lipids B) in both groups (n=2); C) only in CMA group (n=39) and D) only in placebo group (n=23) on Day 70 vs Day 0. Heatmap shows log2FC based alterations in the lipids. Asterisks indicate statistical significance based on paired Student’s t test. p< 0.05. Log2FC, log2(fold change).

**Figure S2.**

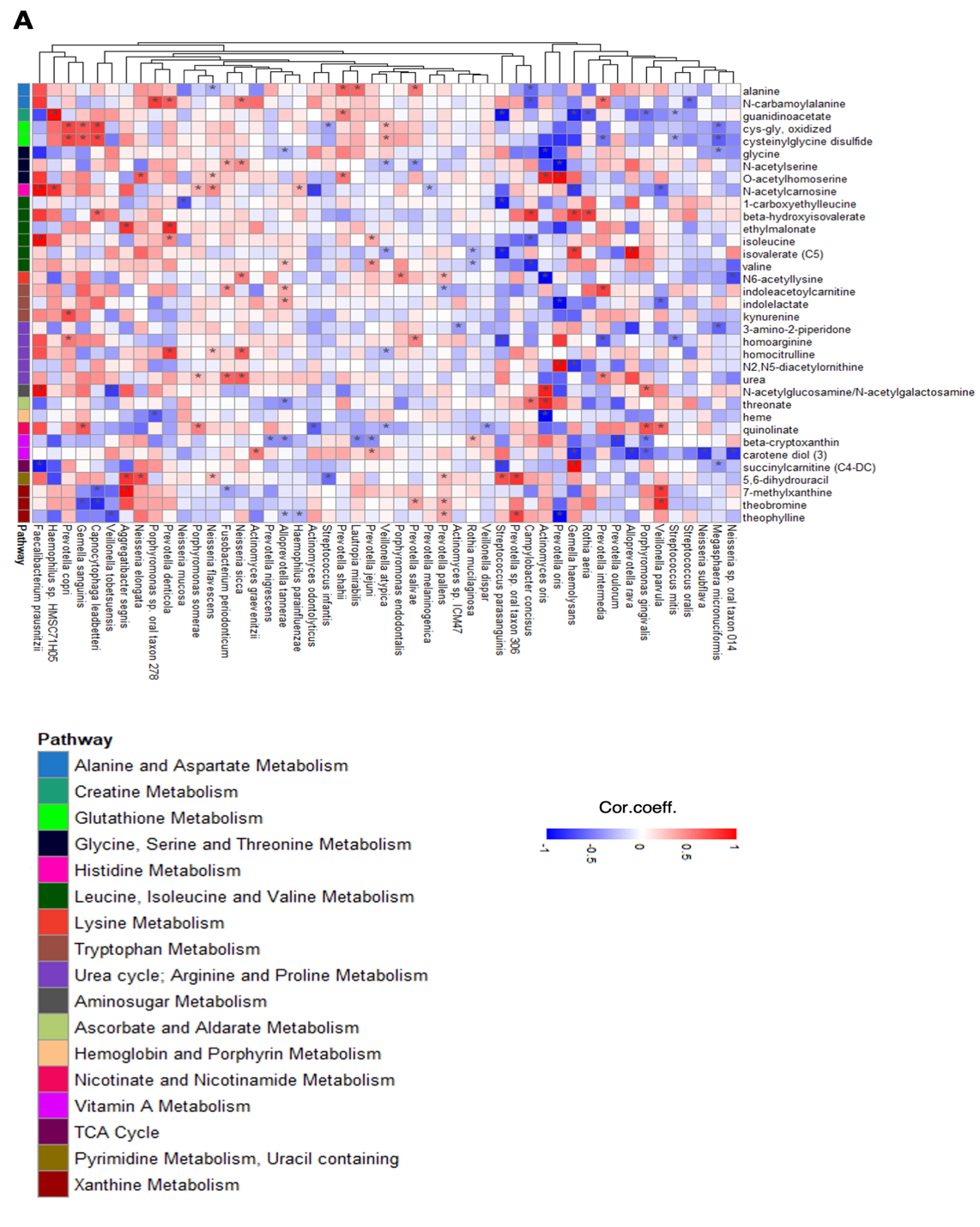

**Figure S2. Associations between the abundance of species in oral microbiome and the plasma level of metabolites (other than lipids)**

Heatmaps show the correlations between the plasma levels of metabolites (other than lipids) and the abundance of the species in oral microbiome. Asterisks indicate statistical significance based on Spearman correlation analysis. p < 0.05; Cor.Coeff: Correlation coefficient

**Figure S3.**

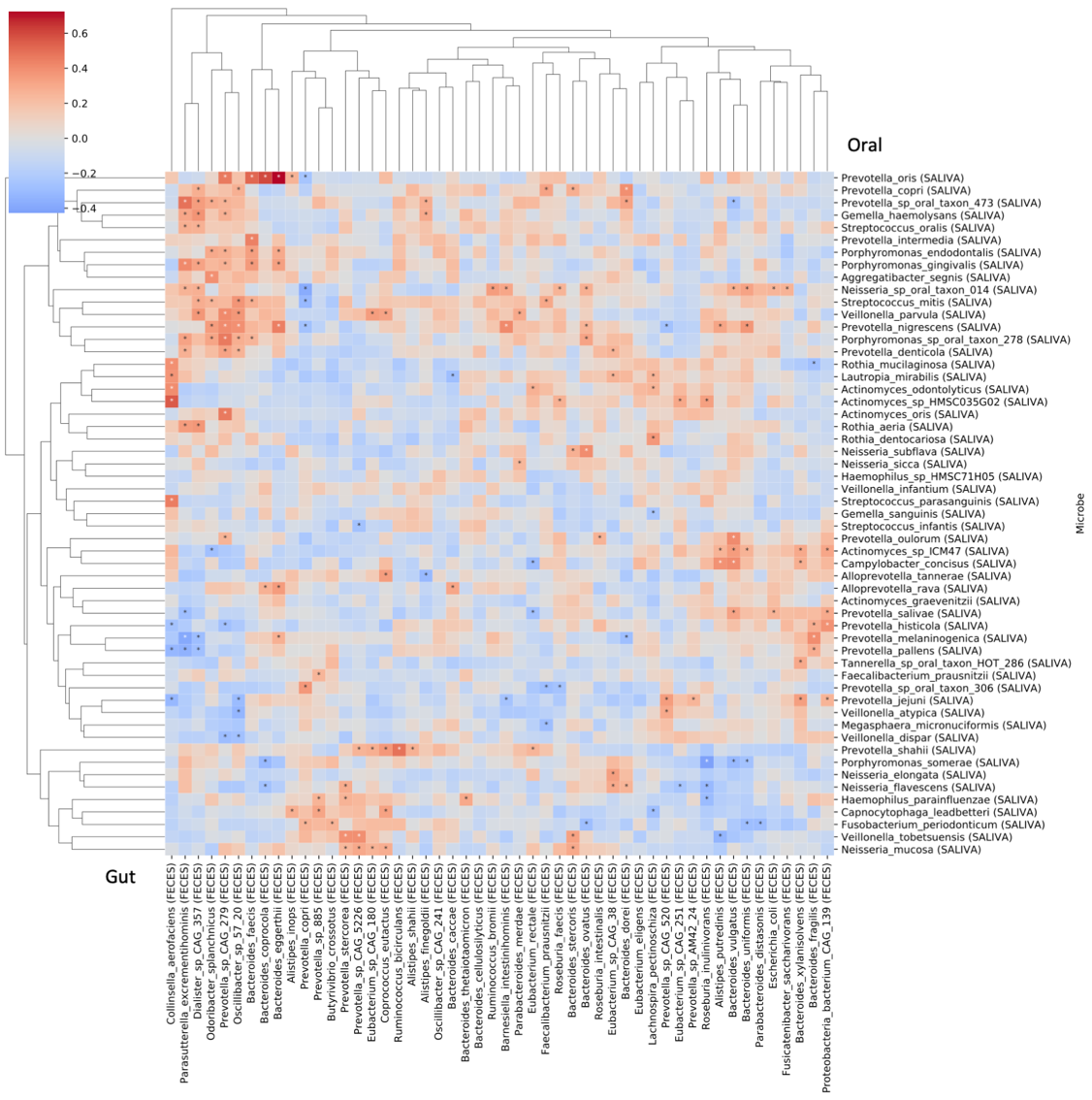

**Figure S3 Associations between the abundance of species in oral microbiome and gut microbiome**

Heatmaps show the correlations between the abundance of the species in oral microbiome and gut microbiome. Asterisks indicate statistical significance based on Spearman correlation analysis. p < 0.05; Cor.Coeff: Correlation coefficient

**Figure S4.**

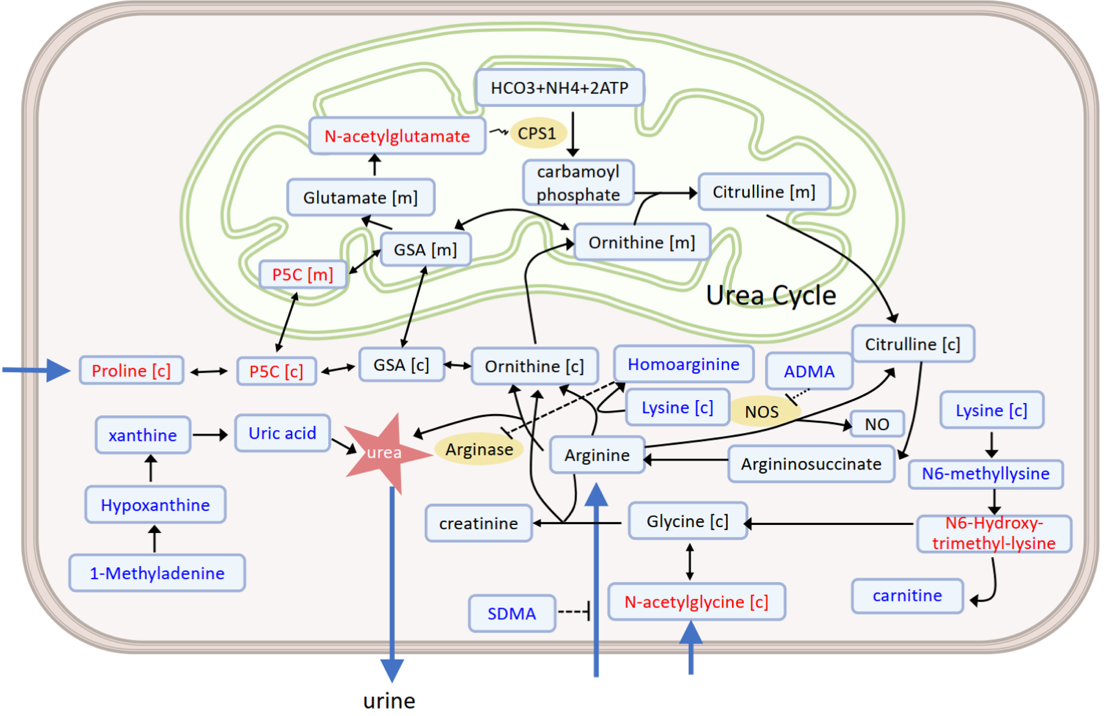

**Figure S4.** **CMA Decreases Uric Acid Plasma Levels**

The uric acid level is significantly decreased on Day 70 vs Day 0 in the CMA group. Changes in metabolites or intermediates in key metabolic reactions are highlighted: Red and blue indicates the metabolite that was significantly increased and decreased, respectively, in the CMA group vs the placebo group on Day 70. Letters in brackets indicate subcellular compartments: [c], cytosol; [m] mitochondria. Arrows and dashed arrow link direct and indirect interactions, respectively. Black dashed line ending in a bar indicates a reaction inhibited by a certain metabolite. For example, asymmetric dimethylarginine (ADMA) is a potent inhibitor of nitric oxide synthase (NOS), whereas symmetric dimethylarginine (SDMA) inhibits nitric oxide (NO) production by inhibiting cellular uptake of arginine. CPS1 is regulated allosterically by N-acetyl glutamate, indicated by paired wavy lines. P5C, 1-pyrroline-5-carboxylate; GSA, L-glutamate 5-semialdehyde; CPS1, carbamoyl phosphate synthase 1. The blue arrows indicated that N-acetylglycine and proline are imported from extracellular, and the urea is secreted.
