## Supplementary_ for "Combined Metabolic Activators Reduces Liver Fat in Nonalcoholic Fatty Liver Disease Patients": 210519_Zeybel et al_E_SupplementaryAppendix.docx

**STUDY PROTOCOL (FINAL, APPROVED)**

### Supplementary Text

Indication:	Nonalcoholic fatty liver disease (NAFLD)

Study agent: Combined metabolic activators (CMA) consisting of serine, L-carnitine tartrate, N-acetylcysteine, and nicotinamide riboside

Study design: Single-blind, randomized, placebo-controlled Phase 2 trial

Treatment arm: At home-treatment with CMA

#### Primary Objective

The primary objective was to assess the difference in the amount of liver fat between subjects treated for 10 weeks with CMA or placebo, as quantified by the proton density fat fraction determined by magnetic resonance imaging (MRI-PDFF)

#### Secondary Objectives

The secondary objectives were (1) to determine the change in liver fat amount as measured by MRI-PDFF within the same groups; (2) to assess the tolerability and safety profile of co-factor supplementation; (3) and to examine additional efficacy variables, including biomarker, lipidomic and metabolomic analysis.

#### Number of Subjects

The protocol called for enrollment of 45 overweight or obese patients with NAFLD were randomized on a 2:1 basis to the CMA or placebo. However, we recruited 32 overweight or obese patients with NAFLD due to COVID-19 and it has not affected the power of the study.

#### Main Inclusion Criteria

1.	Men and women (18–70 years old)

2.	Body mass index >27 kg/m^2^

3.	Triglycerides ≤354 mg/dl and low-density lipoprotein cholesterol ≤175 mg/dl

4.	No history of medication use for hepatic steatosis

5.	Increased liver fat (>5.5%)

#### Main Exclusion Criteria

1.	Inability or unwillingness to give written informed consent

2.	Systolic blood pressure >160 mm Hg and/or diastolic blood pressure >105 mm Hg

3.	Type 1 or type 2 diabetes

4.	Chronic liver disease other than NAFLD (i.e., chronic infection with hepatitis C or hepatitis B, autoimmune hepatitis, primary biliary cirrhosis, primary sclerosing cholangitis, Wilson’s disease, alpha-1 antitrypsin deficiency)

5.	Previous gastric or small bowel surgery

6.	Active gastric ulcer

7.	Inflammatory bowel disease

8.	Alanine aminotransferase or aspartate aminotransferase >3× the upper limit of normal

9.	Cirrhosis detected by transient elastography or other imaging techniques

10.	Diarrhea (more than 2 stools per day) within 7 days before enrollment

11.	Chronic kidney disease (i.e., estimated glomerular filtration rate <60 ml/min/1.73 m^2^

12.	Significant cardiovascular comorbidity (i.e., heart failure, documented coronary artery disease, valvular heart disease)

13.	Active bronchial asthma

14.	Phenylketonuria (contraindicated for N-acetylcysteine)

15.	Histamine intolerance

16.	Clinically significant thyroid-stimulating hormone level outside the normal range (0.04–6 mU/L)

17.	Known allergy to substances used in the study

18.	Concomitant medication use

a. Lipid-lowering drugs within 3 months

b. Oral antidiabetics for insulin resistance of obesity (e.g., metformin, liraglutide) within 3 months

c. Thiazide diuretics with a dose >25 mg/d

d. Postmenopausal estrogen therapy

e. Any medication acting on nuclear hormone receptors or inducing CYP P450

f. Self-administration of dietary supplements (e.g., any vitamins, omega-3 products, or plant stanol/sterol products) within 1 month

g. Treatment with medications known to cause fatty liver disease (e.g., atypical neuroleptics, tetracycline, methotrexate, or tamoxifen)

h. Use of an antimicrobial agent in the 4 weeks before randomization

19.	Smoking (>10 cigarettes/day)

20.	Alcohol consumption >192 g/week for men and >128 g/week for women

21.	Patients deemed inappropriate for this study for any reason (e.g., unable to undergo MRI study, noncompliance)

22.	Subjects with PNPLA3 I148M (homozygous for I148M)

23.	Women who were pregnant, planning pregnancy, or breast-feeding

24.	Women of childbearing potential not protected by effective birth control

25.	Active participation in another clinical study

#### Dosage and duration of therapy

Each dose of CMA contained L-carnitine tartrate, 3.73 g; N-acetylcysteine, 2.55 g; nicotinamide riboside, 1 g; and serine, 12.35 g. Patients took one dose daily for 14 days (after dinner) and two doses for the next 56 days (after breakfast and dinner). The total duration of the therapy was 10 weeks.

Investigational Products

CMA and placebo were packaged as individual doses in powdered form in identical screwtop, 60-ml HDPE plastic bottles. Strawberry flavor is added to both CMA and placebo. Each dose was dissolved in 200 ml of tap water before use.

#### Study design

The study comprised a screening period and 3 visits

**Screening.** Between 4 weeks and 1 day before the start of the study, the participants received oral information on the study protocol, including genetic and imaging studies, and signed the informed consent form. Liver fat content and stiffness were estimated by CAP and transient elastography. Clinical and physical examinations were done, blood samples were obtained (for biochemistry, pregnancy test, and genetic studies), body composition was analyzed by noninvasive bioelectrical impedance analysis, an ECG was done, and blood and urine samples were obtained for routine analysis at Koç University Hospital.

The participants underwent MR imaging (MRI) to determine liver fat content. Plasma samples were taken for advanced proteomics and metabolomics analysis in Sweden and USA, respectively. Those with ultrasound-positive liver steatosis and a genetically eligible profile came to Visit 1.

**Visit 1 (Day 0):** Study subjects with increased liver fat (>5.5%) determined by MRI-PDFF were invited back to the clinic for randomization to the active treatment and placebo groups. They were asked to take two doses at the hospital and were observed for side effects. Patients who tolerated the study agents continued to take them as described above.

**Visit 2 (Day 14±3):** The subjects returned to the study center for complete evaluation, including analysis of body composition and recording of adverse events. Liver fat content was determined by MRI-PDFF. Biochemical, proteomic, and metabolomic analyses were repeated, including ECG, as at Visit 1. After Visit 2, subjects started to take 2 doses of supplements daily.

**Visit 3 (Day 70±3):** The subjects returned for a final evaluation consisting of clinical and physical examinations, ECG, analysis of body composition analysis, recording of adverse events recording, determination of liver fat, by MRI-PDFF, and biochemical, proteomic, and metabolomic analyses. After Visit 3, subjects stopped taking their supplements.

#### Study duration

The active treatment duration was 10 weeks for each subject. The total study duration was 14 months.

#### Efficacy evaluation

The primary analysis was to determine the difference in liver fat content between the placebo and the treatment arms, as quantified by MR-PDFF, at Visit 3. The secondary analysis assessed repeated measurements of liver fat content, as quantified by MR-PDFF, within and between groups and between Visits 1 and 3. The tertiary analysis assessed panels of metabolic and cardiovascular biomarkers.

#### Safety evaluation

Safety was assessed by monitoring adverse events, physical examination, vital signs measurements, and clinical laboratory tests. Patients were monitored for adverse events by weekly phone contacts.

|  | **Visit** | | | |
| --- | --- | --- | --- | --- |
| **Visits** | **Screening** | **Visit 1** | **Visit 2** | **Visit 3** |
| **Days** | **Day 0** | **Day 0** | **Day 14** | **Day 70** |
| Informed consent | X |  |  |  |
| Demographic data | X |  |  |  |
| Surgical & medical history | X |  |  |  |
| Physical examination^1^ and diagnosis | X | X | X | X |
| **Inclusion/exclusion criteria** | X |  |  |  |
| **Laboratory tests** |  |  |  |  |
| Blood sample collection^2^ | X | X | X | X |
| Body composition^3^ | X | X | X | X |
| DNA analysis^4^ | X |  |  |  |
| Serum metabolomics analysis^5^ |  | X | X | X |
| **Radiological assessments^6^** |  |  |  |  |
| Transient elastography | X |  |  |  |
| MRI | X | X | X | X |
| **Efficacy and safety evaluation** |  |  |  |  |
| Laboratory safety parameters ^7^ | X | X | X | X |
| ECG evaluation | X | X | X | X |
| Pregnancy test^8^ | X | X | X | X |
| Urine test^9^ | X | X | X | X |
| **Study drug administration** |  |  |  |  |
| Randomization ^10^ | X |  |  |  |
| Drug administration^11,12^ |  | X | X | X |
| Monitoring of compliance ^13^ |  | X | X | X |
| Adverse events ^14^ |  | X | X | X |

^1^Physical examination included body weight, height, body mass index, vital signs, and measurement of waist and hip circumferences.

^2^At the screening visit, whole-blood samples were collected for DNA extraction. At visits 1, 2 and 3, whole-blood samples were collected for -omics analysis and assessment of safety variables.

^3^Body composition was analyzed by noninvasive bioelectrical impedance analysis to calculate estimated total body water, body fat, and amount of lean tissue.

^4^DNA was extracted from whole blood to identify common genotypes (PNPLA3) that confer susceptibility to NAFLD due to abnormal hepatic lipid handling.

^5^Serum metabolic and proteomic analyses included generation of untargeted metabolomics and proteomic data in Sweden.

^6^At the screening visit, the liver was assessed by transient elastography, and liver fat content was assessed by MRI-PDFF. At Visits 1, 2, and 3, liver fat content was estimated by MRI-PDFF.

^7^Laboratory safety variables included complete blood count (blood cells, hemoglobin), alkaline phosphatase, alanine aminotransferase, aspartate aminotransferase, gamma-glutamyl transferase, bilirubin, albumin, creatinine kinase, total cholesterol, high-density lipoprotein cholesterol, low-density lipoprotein cholesterol, triglycerides, creatinine, urea, urate, glucose, sodium, potassium, insulin, HbA1C (Visit 2 only), and thyroid-stimulating hormone.

^8^ A pregnancy test was done only in women of childbearing potential.

^9^ A dipstick test was done to assess albuminuria.

^10^Subjects with increased liver fat (>5.5%) determined by MRI-PDFF at screening were offered to be enrolled in the study and invited back to the clinic for randomization and to take the first dose at the hospital.

^11^Subjects took one dose just after dinner between Day 0 and Day 14.

^12^Subjects took two equal doses after breakfast and dinner between Day 15 and Day 70.

^13^Compliance and adverse events were assessed by weekly phone contact with the study nurse.

^14^Adverse events and serious adverse events were monitored continuously and reported in Case Report Forms.
